# Dual site proteomic analyses reveal potential drug targets for cardiovascular disease

**DOI:** 10.1101/2024.07.27.24311104

**Authors:** Christopher Aldous Oldnall, Julian Ng-Kee-Kwong, Jimi Wills, Anne Richmond, Tim Regan, Sara Clohisey Hendry, Archie Campbell, J. Kenneth Baillie, Alex von Kriegsheim, Chris Haley, Ava Khamseh, Sjoerd Viktor Beentjes, Andrew D. Bretherick

## Abstract

**Background:** While genome-wide association studies (GWAS) hold great promise for unravelling disease pathophysiology, the translation of disease-associated genetic loci into clinically actionable information remains a challenge. Mendelian randomisation (MR), using expressed proteins as exposures and disease as an outcome, stands as a powerful analytical approach for leveraging GWAS data to identify potential drug-targets—at scale—in a data-driven manner. Cardiovascular disease (CVD) is a major health burden worldwide, and therefore is an important outcome for which to establish and prioritise potential therapeutic targets.

**Methods:** In this study, we utilised generalised summary-data-based MR (GSMR) with novel mass-spectrometry-based isoform-specific protein groups measured from peripheral-blood mononuclear cells (PBMCs) obtained from Generation Scotland and antibody-based plasma protein measures from UK Biobank as exposures, and two CVD and three CVD-related risk-factors from UK Biobank as outcomes. To ensure transparent reporting of our MR findings we adhere to the STROBE-MR guidelines. Further, we used colocalisation to assess support for a shared causal variant between the proteins and the disease outcomes providing further evidence supporting a causal link.

**Results:** We evaluate expression of 5,114 isoform-specific protein groups in PBMCs from 736 individuals. GSMR analysis, using this data, found five putative causal proteins across three of the CVD/CVD-related risk-factors with support for all five by colocalisation analysis. Within the plasma GSMR analysis, 92 putative causal proteins were identified, with 59 supported by colocalisation. In addition, we go on to examine enrichment amongst the results and find enrichment of pathways which relate to cholesterol metabolism. COMT was the only protein identified as significant by GSMR in both sets of data, demonstrating a consistent direction of effect in both Type II Diabetes and Essential Hypertension.

**Discussion:** This study identifies a number of proteins and pathways that may be involved in CVD pathogenesis. It also demonstrates the importance of the location of protein measurement and the methods by which it is quantified. Our research contributes to ongoing efforts to bridge the gap between genotype and phenotype.

**Author Summary:** Cardiovascular disease (CVD) is one of the leading causes of illness and death worldwide. While genetics plays a significant role in disease mechanisms, connecting genome-wide association study findings to specific proteins that could serve as drug targets remains a challenge. Peripheral blood mononuclear cells (PBMCs) are a group of immune cells that includes T-cells, B-cells, monocytes, and natural killer cells. Such cell-types are intricately linked to many inflammatory processes, of which atherosclerosis is one. In this study, we analysed the effect of protein levels in both plasma, as measured by an affinity-based assay, and PBMCs, as measured by mass-spectrometry, on CVD and its risk-factors in large population cohorts. We used a technique called Mendelian randomisation, which mimics a randomised trial by using naturally inherited genetic differences, to test whether certain proteins influence disease risk. Our study identified several promising protein targets, including some that were only identified in plasma, or in PBMCs. This highlights the importance of the tissue- or cell-type in which proteins are measured, as well as the importance of considering the measurement technique used, and genetic variant selection when interpreting the results of Mendelian Randomisation studies. These findings provide a shortlist of proteins for future research and potential therapeutic development, moving us a step closer to translating genetic insights into effective treatments for CVD.

## 1 Introduction

Cardiovascular disease (CVD) is of clear global importance [1–3]. With respect to its genetics, it is a complex-trait, underpinned by multiple genetic variants. Genome-wide association studies (GWAS) have emerged as a powerful tool for uncovering the intricate genetic architecture of many diseases, including CVD. However, a substantial portion of disease-associated variation resides outside the coding sequence of genes and the translation of GWAS findings into actionable clinical insight remains a significant challenge [4].

Here we aim to identify proteins that are on the causal pathway to CVD, and its risk-factors. By combining a novel data set of protein-quantitative trait loci (pQTLs) identified by mass-spectrometry in peripheral-blood mononuclear cells (PBMCs) from Generation Scotland (GS) [5], pQTLs in plasma from the UK Biobank (UKB) [6], and GWAS of CVD and CVD-related risk-factors, we identify 96 unique putative drug-targets by employing a two-sample Mendelian randomisation (MR) approach. MR identifies naturally occurring subgroups with genetically determined high, or low, values of an exposure (e.g., a protein) and tests for an association between these groups and outcome (e.g., CVD). Since the set of genetic variants inherited by an individual is (approximately) random, MR is akin to a ‘natural’ randomised controlled trial.

Under such circumstances, the finding that a genetic variant is significantly associated with the risk-factor under investigation, which may be a particular protein, while also being associated with the disease, suggests a potential causal role for that protein in that disease. It should be noted that the following three assumptions must hold to ensure the validity of such an analysis: (i) the genetic variant must be strongly associated with the protein of interest, (ii) it must be independent of any confounding of the protein and phenotype, and (iii) it must influence the disease solely through its effect on the protein in question, i.e., there is no horizontal pleiotropy [7]. When a genetic variant satisfies these three assumptions it is called a valid instrumental variable (IV).

Summary data-based Mendelian Randomization (SMR) and generalised SMR (GSMR) have emerged as MR techniques that integrate data from independent studies [8, 9]. These methods harness GWAS summary-level data, thereby allowing analyses to be completed where the exposure and outcome have been measured in two different cohorts sampled from the same underlying population. Additionally, the GSMR pipeline incorporates outlier analysis (termed HEIDI), which attempts to identify and remove potential pleiotropic effects within the single nucleotide polymorphism (SNP) set. This is done to ensure that all included genetic variants are valid IVs. Colocalisation of the exposure and outcome is also used here, in conjunction with the MR analyses, in order to provide further support for potential therapeutic targets [10]. Bayesian testing for colocalisation using the software ‘coloc’ allows for intuitive interpretation of posterior probabilities aligning with different colocalisation possibilities [11].

Previous studies have employed RNA expression (eQTLs) and pQTLs in MR analyses with great success, identifying putative drug-targets (e.g., see [12–14]). While substantial literature exists on statistical methodology related to IV analysis [15], using MR analysis to assess the functional significance of genes at disease-associated loci presents its own set of challenges. Of particular relevance here, for practical reasons, it is often difficult to measure RNA/protein abundance in a disease-relevant tissue/cell-type. Indeed, most previous pQTL studies have been conducted using proteins measured in plasma [6, 16, 17]. Given the lack of available cellular pQTLs, there has been limited exploration of MR using cell-based measures of proteins abundance to date. PBMCs include many of the fundamental cells of the immune system, including B-cells, T-cells, monocytes, and natural killer (NK) cells. Such cell-types are intricately linked to many inflammatory processes, of which atherosclerosis is one. Proteomics analysis of PBMCs complements plasma protein measurement by providing insight into cellular pathways as well as immune-cell specific protein abundance.

In this study, we present the results of our investigations using GSMR and colocalisation techniques to establish causal links between proteomic data, from both novel cellular pQTL data from PBMCs (GS) and plasma (UKB), and CVD/CVD-related risk-factors from UKB. Here we compare results simultaneously for proteomic associations with CVD/CVD-related risk-factors in both sets of protein data. Throughout this study, we address the items of the STROBE-MR guidelines [18] to ensure transparency and clarity in the presentation and interpretation of our MR findings.

## 2 Results

### 2.1 Cellular pQTL analysis reveals 127 significant SNP-protein expression associations

The majority of previous pQTL data have been measured in plasma. We performed genome-wide association of the cellular protein abundance for 5,114 isoform-specific protein groups in PBMCs from GS. At an uncorrected genome-wide significance threshold (*p <* 5 × 10*^−^*^8^), 663 isoform-specific protein groups were identified to have at least one SNP, associated with protein abundance. After applying a Bonferroni correction, 127 isoform-specific protein groups were identified which had at least one significantly associated SNP (*p <* 5 × 10*^−^*^8^*/*5114).

Subsequent analyses focused on proteins with at least one cis-pQTL (±1Mb from the gene encoding the protein) significant at a Bonferroni corrected genome-wide significance threshold (*p <* 5 × 10*^−^*^8^*/*5114) [19–21]. This approach narrowed our focus to 99 isoform-specific protein groups for GSMR (Fig. 2A). Fig. 1 displays, as exemplars, Manhattan plots for the GWAS results of six proteins which are of subsequent interest in this paper: COMT, FN3K, FN3KRP, HLA-B, HLA-DRA, and HLA-DRB1.

**Fig. 1:**
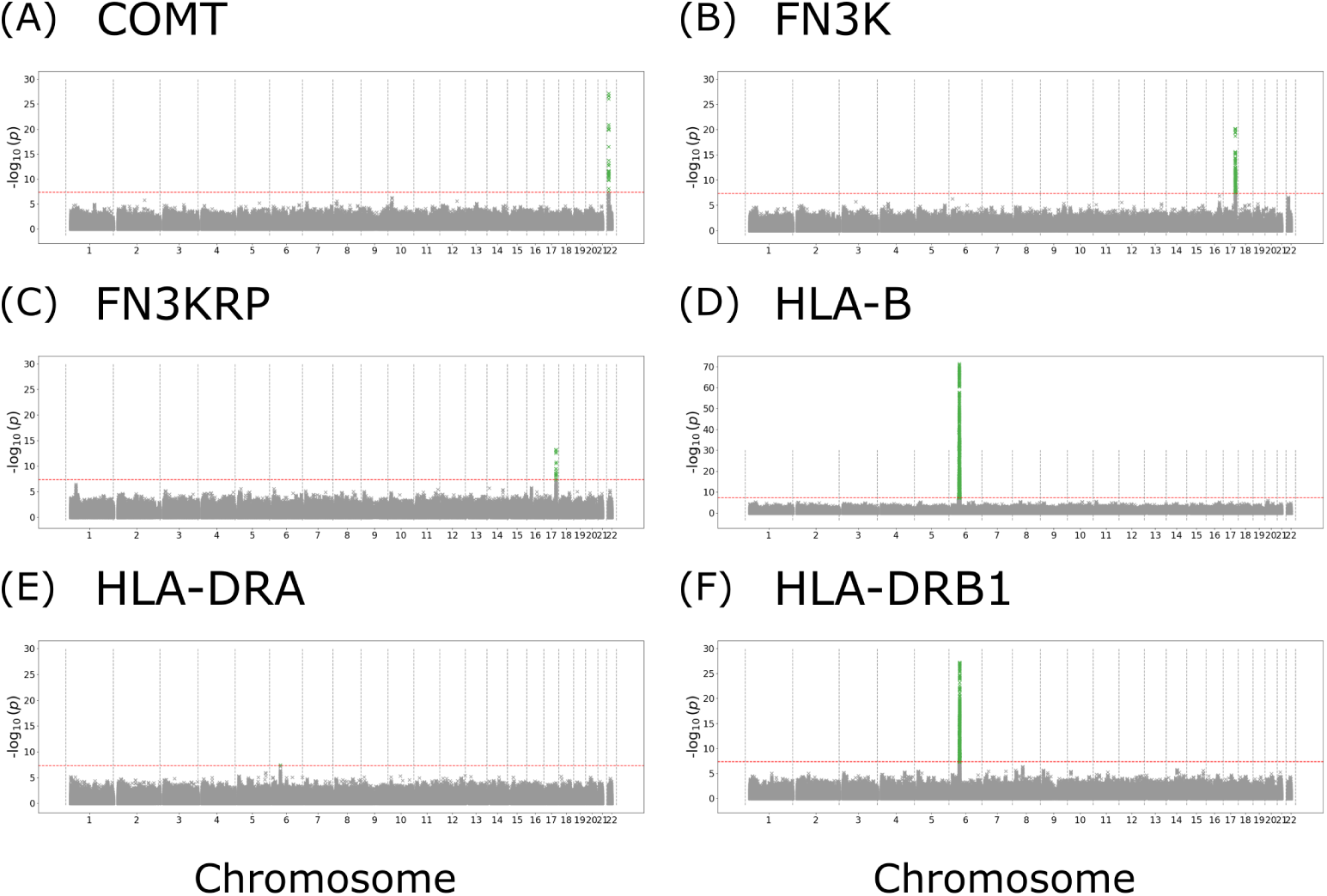
Genome-wide SNP associations with PBMC protein abundance. Manhattan plots depicting − log_10_(*p*) of SNPs across all chromosomes for **(A)** COMT, **(B)** FN3K, **(C)** FN3KRP, **(D)** HLA-B, **(E)** HLA-DRA, and **(F)** HLA-DRB1 proteins. SNPs associated with the protein below the genome-wide significance threshold (*p <* 5 × 10*^−^*^8^) are denoted in green. The red dashed line is at − log_10_(5 × 10*^−^*^8^)) corresponding to the genome-wide significance level per protein.

**Fig. 2:**
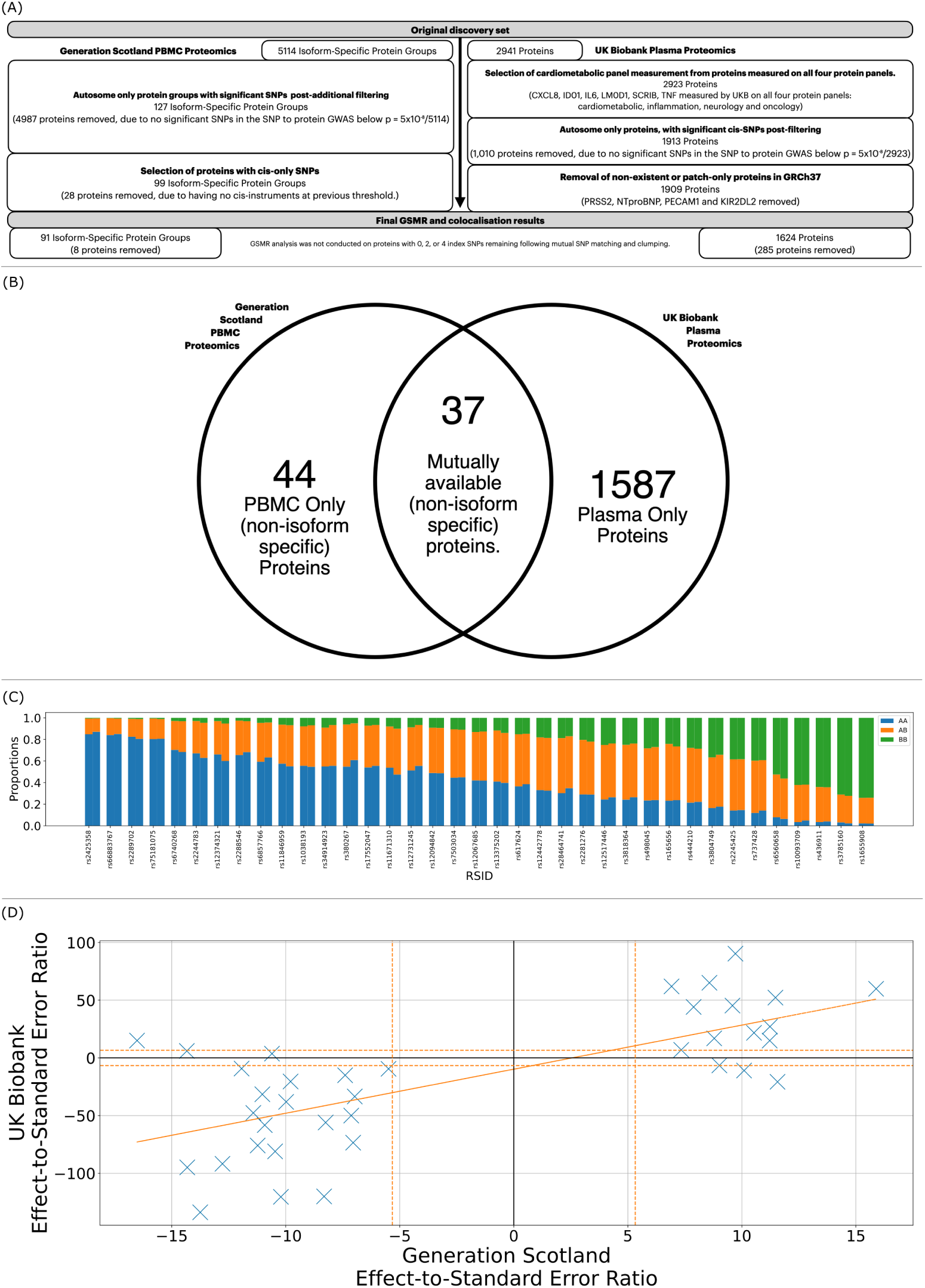
Filtering and comparison of the PBMC and plasma proteomic data and populations. **(A)** Filtering process of both PBMC and plasma measured proteins. Only those proteins which had at least one associated genetic variant with the expression (at either *p <* 5 × 10*^−^*^8^*/*5114 for the isoform-specific protein groups in PBMCs, or *p <* 5 × 10*^−^*^8^*/*2923 in the plasma measured proteins) were retained. Additional filtering included use of cis-only genetic variants, removal of non-autosomal proteins, and those which did not map to GRCh37 (plasma proteins only). Proteins with either; 0, 2 or 4 index genetic variants remaining, following clumping, did not have GSMR analysis conducted. **(B)** Venn diagram illustrating the overlap between protein measured in PBMCs and plasma. Proteins included in the diagram are those for which GSMR analysis was conducted, are non-isoform-specific and had at least one cis-genetic variant associated with their abundance (Bonferroni correction, *p <* 5 × 10*^−^*^8^*/*5114 for PBMCs or *p <* 5 × 10*^−^*^8^*/*2923 for plasma). **(C)** Comparison of the allele frequencies of the 37 lead SNPs in the PBMC data, where the protein is measured in both PBMC and plasma data, and whose abundance is associated with a cis-SNP at a Bonferroni-corrected genome-wide significance level *p <* 5 × 10*^−^*^8^*/*5114 in the PBMCs. For each of the 37 proteins, the lead SNP with respect to expression of the canonical isoform is selected and its allele frequency reported. The rsID of the lead SNP is denoted on the x-axis, whilst the y-axis displays the proportions of alleles (AA, AB, or BB) for GS and UKB next to one another, i.e., one bar has both the allele frequencies of the same SNP from GS and UKB next to each other. As can be seen, the relative frequencies of alleles are similar between the two cohorts. **(D)** Comparison of effect-to-standard error ratio in the PBMC and plasma GWAS, of the lead SNP with respect to abundance in the PBMCs, for the 37 proteins present in both the PBMC and the plasma data. On the x-axis are the effect-to-standard error ratios for the lead SNP to abundance association in the PBMCs, and on the y-axis are the effect-to-standard error ratios in plasma. The linear correlation is *R* = 0.709 (*p* = 8.904 × 10*^−^*^7^). Three SNPs are significantly associated with the abundance of the same protein in both PBMCs and plasma, but with the opposite direction of effect—these can be found in the upper-left and bottom-right quadrants. This provides evidence that there is a difference between cellular and plasma expression and/or the differing measurement modalities.

Mass-spectrometry measures the abundance of peptide fragments of a protein, and its isoforms, as included in UniProt. If the sequence changes, for example due to missense variation, the detection of the peptide containing the changed residue will be impacted.

We identified 570 modification-specific peptides with at least one SNP that was associated with expression at *p <* 5 × 10*^−^*^8^/5,114, and 541 at *p <* 5 × 10*^−^*^8^/29,639, corresponding to the number of isoform-specific protein group and modification-specific peptide GWAS considered, respectively; of these modification-specific peptides, 39% (218/556) and 40% (210/527) contained a missense variant. Peptides lacking a common missense variant in their sequence are more likely to reflect true expression differences, suggesting the associated SNP has a pronounced effect on protein expression.

### 2.2 Comparison of cellular and plasma pQTLs shows 37 mutual (non-isoform-specific) instrumented proteins

The UK Biobank pharma-proteomics-project (UKB-PPP) provides a set of 2,941 unique proteins [6]. The dataset underwent a filtering process, as outlined in Fig. 2A, and elaborated in the Methods section 4.2. Post-filtering, which included the removal of duplicate measurements, imposing the requirement for proteins to have at least one cis-genetic variant linearly associated with expression after Bonferroni correction (*p <* 5 × 10*^−^*^8^*/*2, 923), and the reconciliation of genetic variants across datasets, we obtained GSMR and colocalisation results for 1,624 proteins. Out of the initial 99 PBMC-measured proteins with at least one cis-genetic variant associated with cellular expression at a Bonferroni corrected threshold (*p <* 5 × 10*^−^*^8^*/*5114), considering only those for which both GSMR and colocalisaton results were obtained and only the non-isoform-specific form of the protein, 37 were also found in plasma and had genetic variants meeting the Bonferroni corrected threshold for plasma expression, as displayed in Fig. 2B.

We assessed the allele frequencies of the 37 lead SNPs (the SNP most significantly associated with protein expression) of the PBMC-measured isoform-specific protein groups that were measured in plasma. The two populations are similar in allele frequencies, see Fig. 2C, giving confidence in the use of GS SNP-protein data in conjunction with UKB genetic variant-outcome data for MR and colocalisation analyses. Furthermore, we examined the effect-to-standard error ratios of these SNPs on expression levels in plasma and PBMCs. A strong linear correlation (*R* = 0.709*, p* = 8.904 × 10*^−^*^7^) was observed, see Fig. 2D. Together, these findings suggest that any subsequent non-concordance between cellular and plasma MR analysis results may be due to expression difference, measurement technique used, or statistical power.

### 2.3 Protein-to-disease

#### 2.3.1 Analyses with PBMC measured proteins identifies five protein targets for CVD/CVD-related risk-factors

There were five isoform-specific protein groups involved in seven significant protein-to-outcome relationships (FDR *<* 5%, Benjamini–Hochberg [22]), identified using GSMR, with only cis-SNPs, for the CVD/CVD related-risk outcomes (see Supplementary Table 1 for disease prevalence/incidence and population demographics). Each of these isoform-specific protein groups corresponded to either a single protein, or isoforms of the same protein. Angina to one protein, type II diabetes to five proteins and essential hypertension to two proteins. No significant relationships were identified for the outcomes ‘heart attack’ or ‘high cholesterol’. Colocalisation analyses showed support for a range of these same proteins – the probability, H4, that the protein and outcome share a single causal variant. The significant results after multiple hypothesis correction (FDR *<* 0.05, Benjamini–Hochberg) are presented in Fig. 3 and Table 1. Full PBMC results are provided in Supplementary Table 2.

**Fig. 3:**
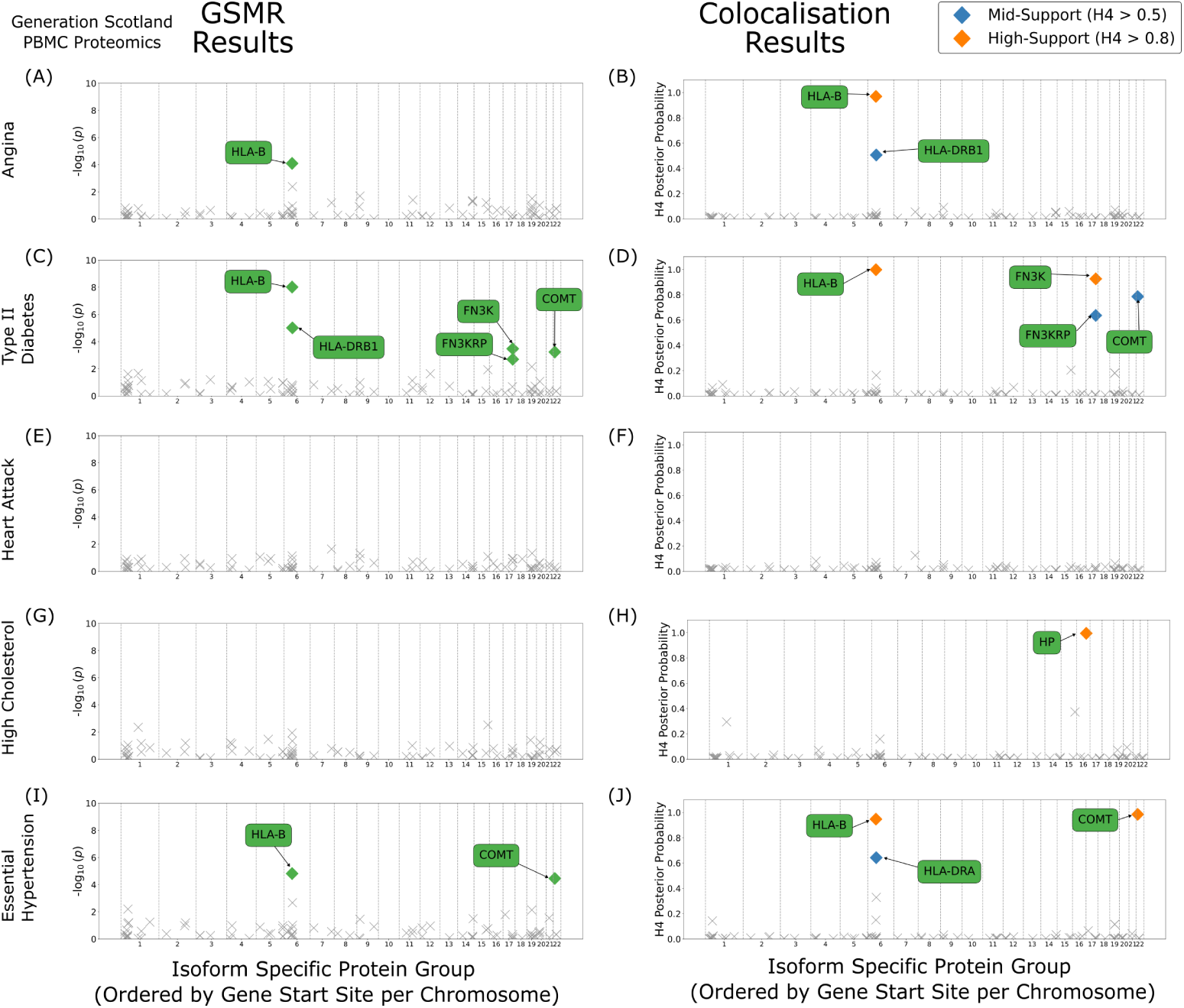
PBMC GSMR and colocalisation results. Left: GSMR results obtained by combining the SNP-isoform-specific protein group associations from the Generation Scotland PBMC proteomics and the SNP-outcome associations from UKB phenotypes. For each phenotype, GSMR results are labelled when they are significant after multiple hypothesis testing correction (FDR *<* 0.05, Benjamini–Hochberg). **Right:** Colocalisation results estimating posterior probability support for the *H*_4_ hypothesis that both the exposure and outcome share a common causal SNP for the corresponding trait. The posterior probability for *H*_4_ is coloured coded: 0.5 – 0.8 (blue), or *>* 0.8 (orange); a greater number indicating greater support for the hypothesis. Isoform-specific protein groups are ordered by the genomic position of their transcription start site. Traits are as follows: **(A-B)** Angina, **(C-D)** Type II Diabetes, **(E-F)** Heart Attack, **(G-H)** High Cholesterol, **(I-J)** Essential Hypertension.

**Table 1:**
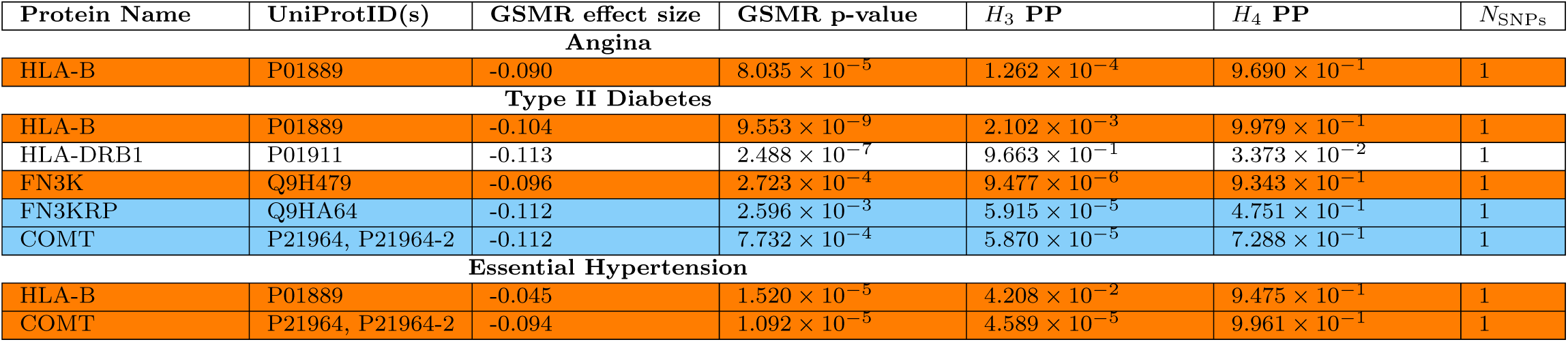
PBMC measured isoform-specific protein group analysis. The table is divided into sections by the CVD/CVD risk-related trait, providing results that are significant in the GSMR analysis. Protein name along with the isoform-specific UniProtID(s) are provided in the first two columns. Additionally, the effect size and p-value from the GSMR procedure and the *H*_3_ and *H*_4_ posterior probabilities (PP) from the colocalisation are given. *H*_3_ corresponds to the hypothesis that both the exposure and outcome have an associated SNP, but a different causal SNP in each case. The number of SNPs used in the GSMR are reported in the final column, ‘*N*_SNPs_. Results are highlighted by H4 colocalisation posterior probability: the probability that both the exposure and outcome share a common causal SNP. Blue corresponds to mid-support, 0.5 *< H*4 *<* 0.8, and orange corresponds to high-support, H4 *>* 0.8).

FN3K and FN3KRP are causally implicated in type II diabetes using GSMR, with high support (H4 *>* 0.8) from colocalisation for FN3K, prioritising them as putative drug targets for type II diabetes. Additionally, we see COMT (catechol O-methyltransferase) linked to both type II diabetes and essential hypertension with mid and high range colocalisation support, respectively. Two HLA-related proteins are also significant (FDR *<* 0.05, Benjamini–Hochberg) across three traits. HLA-B for angina, type II diabetes and essential hypertension as well as HLA-DRB1 for type II diabetes.

#### 2.3.2 UKB plasma protein analysis provides support for 92 targets

GSMR analysis was run on the 1,624 proteins measured in plasma, post-filtering, and the same 5 CVD/CVD risk-related traits as for the isoform-specific protein groups measured in PBMCs, again with only cis-SNPs as instruments. There were 92 unique proteins identified as significant after multiple testing correction across the different outcomes (FDR *<* 0.05, Benjamini–Hochberg): angina (10 proteins), type II diabetes (24 proteins), heart attack (4 proteins), high cholesterol (19 proteins) and essential hypertension (52 proteins). Colocalisation analyses showed high support for a range of proteins to be colocalised with all five outcomes. Apolipoprotein(a) (LPA) was the only protein found as significant (FDR *<* 0.05, Benjamini-Hochberg) in the GSMR results for all 5 CVD/CVD risk-related traits. Results are shown in Fig. 4, annotated proteins are those which were also significant in the PBMC analysis. The full results are provided in Supplementary Table 3.

**Fig. 4:**
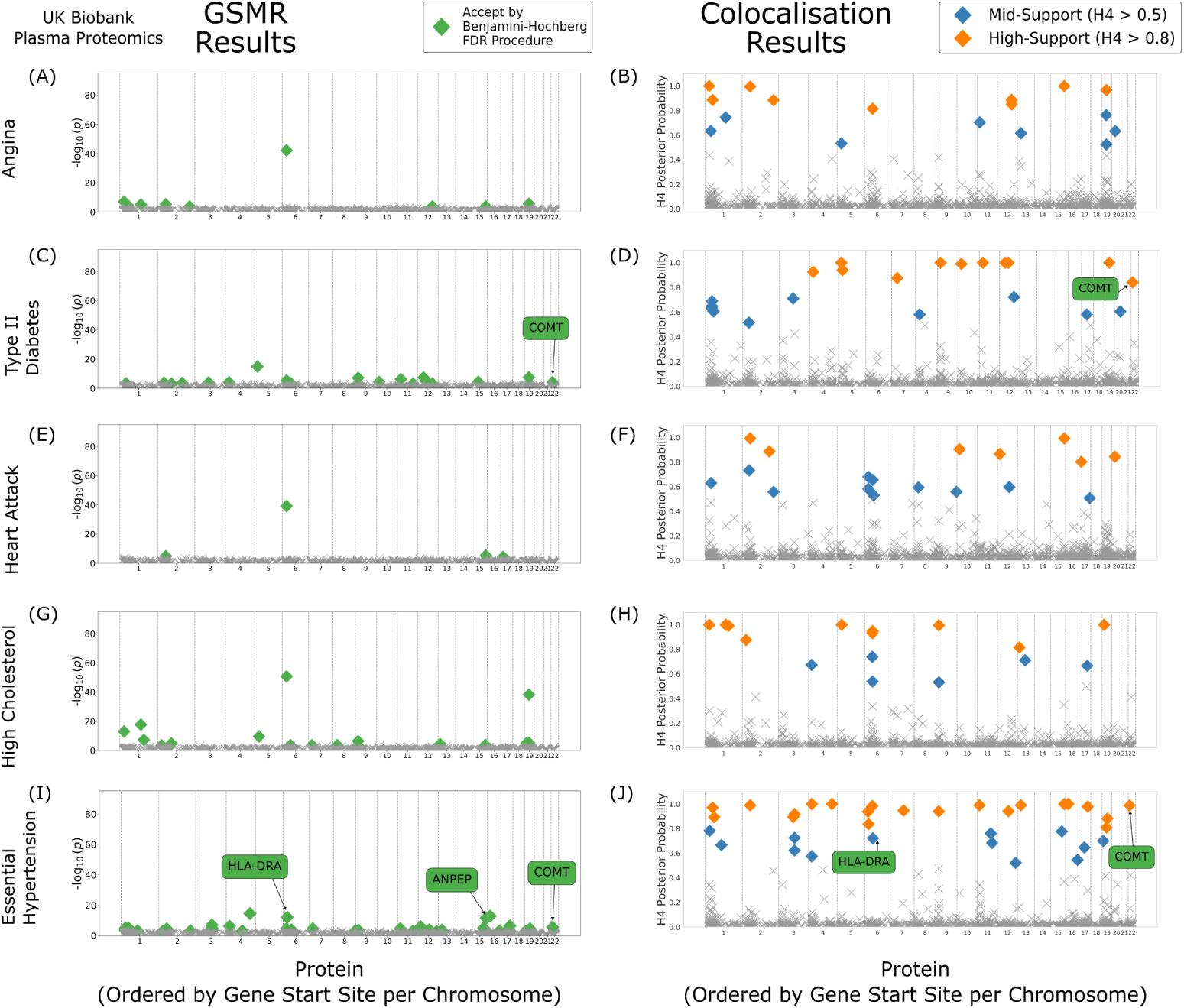
Plasma GSMR and colocalisation results. Left: GSMR results obtained by combining the genetic-variant-protein associations from UKB and the genetic-variant-outcome associations from the UKB phenotypes. **Right:** Colocalisation results estimating posterior probability support for the H4 hypothesis that both the exposure and outcome share a common causal genetic variant for the corresponding trait. The posterior probability for H4 is coloured when between 0.5 and 0.8 (blue) or larger than 0.8 (orange), where a greater number indicates greater support for the hypothesis. Proteins are ordered by the genomic position of their transcription start site. For each trait, GSMR and colocalisation results are labelled if they were significant in the PBMC GSMR results (see Fig. 3) and were also significant after multiple hypothesis correction (FDR *<* 0.05, Benjamini–Hochberg) or had medium/high colocalisation support in the plasma results. Traits are as follows: **(A-B)** Angina, **(C-D)** Type II Diabetes, **(E-F)** Heart Attack, **(G-H)** High Cholesterol, **(I-J)** Essential Hypertension.

Using StringDB [23], we conducted functional enrichment analysis, for the sets of proteins associated with each trait, employing an FDR threshold of 0.05 (Methods 4.7). The final networks of proteins identified by StringDB were of sizes 10, 24, 4, 18, 52 for the traits angina, type II diabetes, heart attack, high cholesterol and essential hypertension respectively. Attrition is due to no matching gene name in StringDB or exclusion of MHC region proteins. The network for angina is provided in Fig. 5, with results highlighted by colocalisation support, and additional network images are provided in Supplementary Figures 1 to 4.

**Fig. 5:**
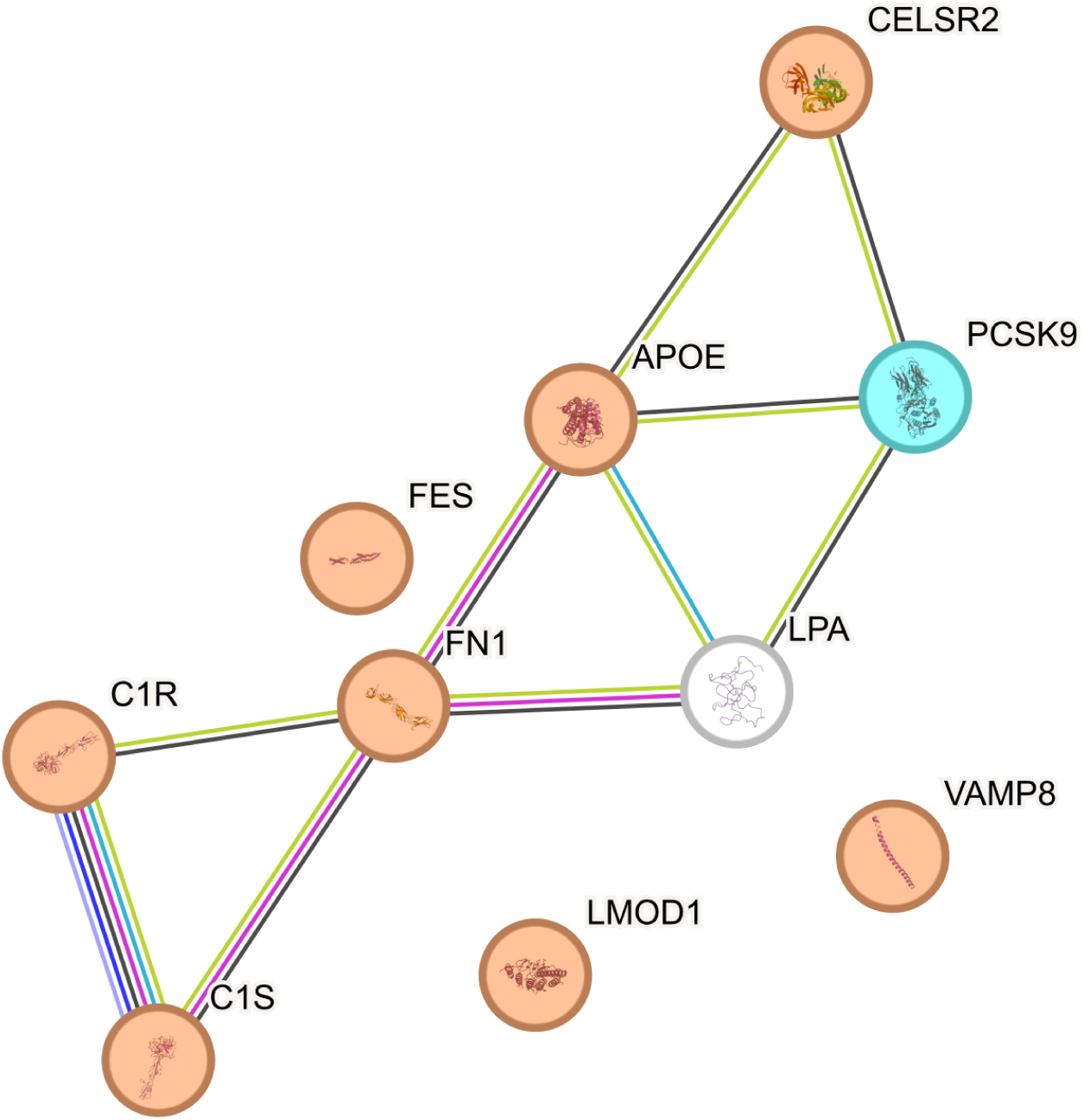
UKB GSMR and colocalisation results network for angina. The String-DB generated network is presented for all non-MHC region proteins (those with a gene starting location outside of chr6:28477797-chr6:33448354) which were found by GSMR to have a significant relationship between the protein and angina (FDR *<* 0.05, Benjamini–Hochberg). Proteins, each represented by a node on the network, are shaded when the H4 colocalisation posterior probability, the hypothesis that the association between the set of genetic variants provided with both the exposure and outcome through one shared genetic variant, is above 0.5. The posterior probability for H4 is coloured when between 0.5 and 0.8 (blue), or larger than 0.8 (orange), where a greater number indicates greater support for the hypothesis. Lines between nodes represent different forms of interaction evidence through: co-occurrence, co-expression, databases, experiments, gene fusion, neighbourhood and text-mining.

Functional enrichment for cholesterol metabolism was found for angina (KEGG, 3/21 proteins; FDR = 0.0309) and high cholesterol (KEGG, 5/21 proteins; FDR = 0.00023). Blood pressure was found to be enriched for essential hypertension (Monarch, 15/151 proteins; FDR = 0.0013). Full functional enrichment analyses from StringDB for all traits are provided in Supplementary Table 4.

### 2.3.3. Concordance in effects of COMT between cellular and plasma results highlights robust association to CVD

Comparing GSMR results obtained using the proteomic data from PBMCs measured by mass-spectrometry, and those measured in plasma using the Olink Explore 3072 PEA platform [6], only COMT was significant (FDR *<* 0.05, Benjamini-Hochberg) in both PBMCs and plasma. An overview of results from both biobanks is presented in Fig. 6.

**Fig. 6:**
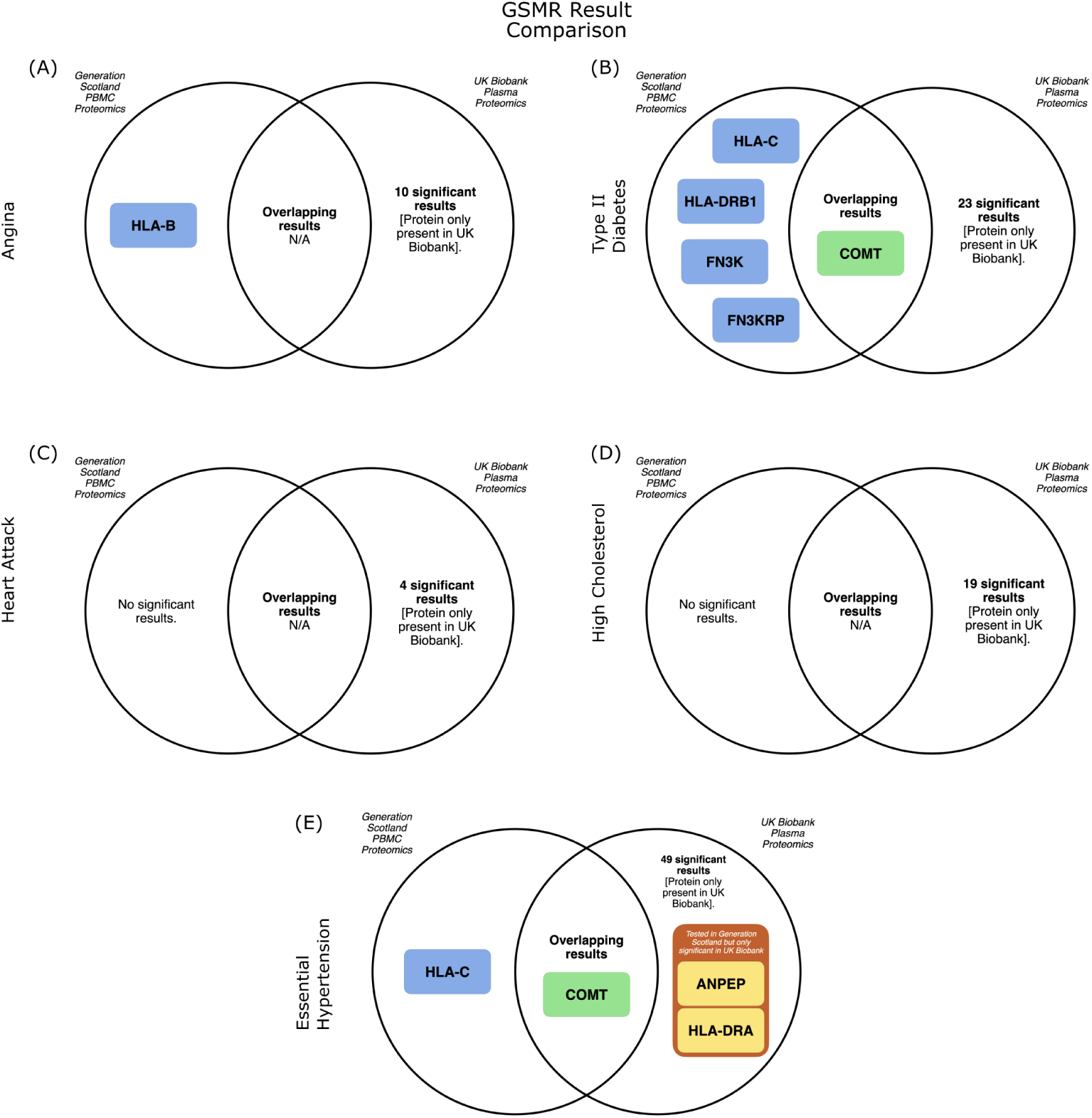
GS/UKB overlap and replication of UKB results in GS. Venn diagrams illustrating the intersection between the PBMC (GS) and plasma (UKB) GSMR significant results (FDR *<* 0.05, Benjamini-Hochberg) in either site, and their overlap. In these diagrams, yellow indicates significance in UKB plasma proteomics but not in GS PBMC proteomics. Blue denotes proteins significant in GS PBMCs but unmeasured in UKB plasma proteomics. Green represents proteins significant in both proteomics sets. The proteins are displayed in relation to various health outcomes: **(A)** angina, **(B)** type II diabetes, **(C)** heart attack, **(D)** high cholesterol, **(E)** essential hypertension.

COMT is observed to have the same estimated direction-of-effect between the two protein measures for type II diabetes and essential hypertension. In the cellular GSMR results, a single SNP was used for COMT; rs165656, a SNP that is in complete LD with rs4680 (*R*^2^ = 1.0, within the 1000 genomes, phase 3 GBR population [24, 25]; https://grch37.ensembl.org/ accessed 05 October 2023), a missense SNP within COMT itself. Despite not being the lead-variant, rs4680 is also strongly associated (*p* = 1.84 × 10*^−^*^27^ compared with rs165656 *p* = 6.70 × 10*^−^*^28^) with expression of COMT in PBMCs. A base change of G to A at rs4680 is associated with a V to M amino acid change. The observed direction-of-effect of a G to A substitution at this SNP is consistent with an observed reduction in the expression of COMT with the unchanged peptide sequence.

Given the nature of mass spectrometry proteomics, it is possible to assess the abundance of the measured modification-specific peptide fragments that have contributed to evaluating the isoform-specific protein group abundance. From COMT (UniProt ID: P21964), there were 17 modification-specific peptides detected. Of these, 10 passed quality control (Methods 4.1.8) and were assessed. However, when applying a Bonferroni correction for the number of peptide GWAS (*p <* 5 × 10*^−^*^8^*/*29, 639), significant association was detected for one only. The sequence of that modification-specific peptide (in this case unmodified) was MVDFAGVK, and is that which contains the expected amino-acid change due to rs4680.

As mass spectrometry proteomics is sensitive to peptide sequence, all else being equal, it is expected that there would be an association between a missense genetic variant and abundance of the peptide containing the associated amino acid substitution. This peptide-level association does not necessarily reflect overall protein abundance: the combined abundance of the protein with and without the amino acid substitution. However, the lead SNP from PBMCs (that most significantly associated with the expression of COMT in the PBMC data) had a consistent direction-of-effect on COMT abundance in plasma, as measured by an affinity-based assay, giving increased confidence in a true effect on abundance. We provide locus zoom plots and a table with details from the corresponding GWAS for the lead SNP in Supplementary Table 5 and Supplementary Figure 5. GSMR Results for all proteins which were instrumented in both biobanks, and significant in at least one, are documented in Table 2.

**Table 2:**
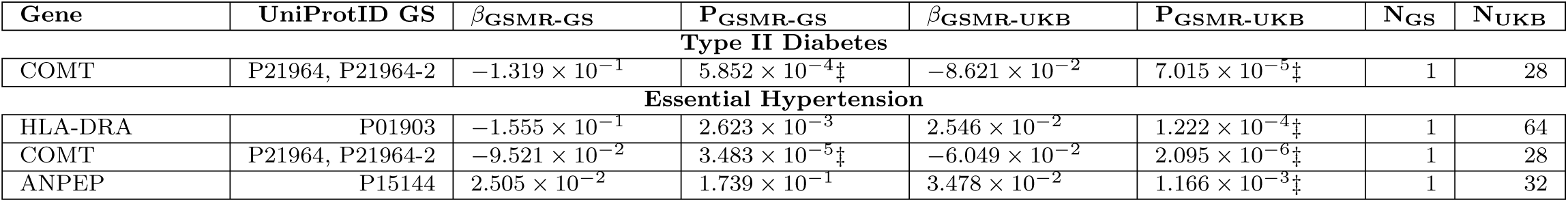
Proteins instrumented in both biobanks, and significant in at least one. . GSMR analysis was conducted on both the PBMC and plasma data sets independently. Shown in the table are the p-values for the GSMR analysis for each gene that was instrumented in both sets of data, and then found to be significant in at least one of the data sets (FDR *<* 0.05, Benjamini–Hochberg). Genes are marked significant, in the corresponding GSMR analyses, with ‡. All genes are accompanied by the UniProtIDs for the isoforms considered in the PBMC data set. The number of instruments used in the GSMR processes are present at the end of the table for each set of data, with a subscript indicating the biobank which the data was obtained from.

## 3 Discussion

Cardiovascular disease (CVD) is a major worldwide cause of mortality and morbidity. In this study we have presented and compared cellular (PBMCs; GS) and plasma (UKB) protein links to five CVD/CVD-related risk-factors, namely angina, type

II diabetes, heart attack, high cholesterol and essential hypertension. We applied generalised summary-data-based MR (GSMR) and Bayesian colocalisation (‘coloc’) to provide a robust list of potential drug-targets for CVD and its risk factors. We examined the causal contribution of 91 proteins in peripheral-blood mononuclear cells (PBMCs) and 1,624 proteins in plasma on cardiovascular disease (heart attack and angina) and CVD-related risk-factors (hypertension, diabetes, and high cholesterol). By measuring proteins in PBMCs from 736 unique individuals – novel data reported here – we are able to comment on protein abundance in a cellular context. Further, given the established role of inflammation in cardiovascular disease, the inclusion of PBMC proteomics in this study allowed us to explore immune-cell protein abundance changes that may underlie CVD traits.

We aimed to identify biological processes shared among those linked to CVD and its associated risk factors through GSMR in plasma. Several enriched protein sets were observed, reflecting key mechanisms underlying disease pathology and treatment. For example, processes related to lipid metabolism (‘Cholesterol metabolism’; KEGG: observed gene count 3; background gene count 21; FDR *<* 0.05) were enriched in angina, aligning with current therapeutic strategies. Additionally, pathways involved in coagulation and inflammation (‘Complement and coagulation cascades’; STRING clusters: observed gene count 7; background gene count 121; FDR *<* 0.05) emerged in relation to CVD, underscoring their role in vascular health.

We have revealed significant putative causal links for five isoform-specific protein groups identified in PBMCs to CVD and its risk factors. Including two paralogs in type II diabetes: FN3K and FN3KRP. In plasma, 92 proteins showed significant associations with one or more CVD-related risk-factors. Notably, LPA was associated across all five outcomes in plasma, highlighting its importance in CVD pathogenesis. Li *et al.* have previously examined the relationship between the UKB-PPP plasma protein data and CVD/CVD risk-related factors, identifying 221 putative proteins [3]. 24 proteins overlapped with our findings, including LPA. The additional 68 targets that, to our knowledge, have not been previously identified in the plasma data set but that we identified – including COMT – are likely due to a range of factors, including, differing genetic-variant-instrument selection criteria, differing CVD/CVD risk-related outcome trait selection and processing, and differing MR analysis methods. COMT was identified as significantly associated with both type II diabetes and essential hypertension in both PBMCs and plasma, with concordant direction-of-effect across tissues, supporting a robust and consistent association.

As with all MR studies, the validity of the conclusions drawn depends on the validity of the underlying assumptions (see Introduction). In order to ensure that the genetic variant (instrument) affects the protein (exposure), we filtered potential instruments based upon the observed strength of this association as part of our analysis pipeline. For UKB data, we additionally ensured that outcomes (diagnosis of disease) were measured after the measurement of the protein (exposure) having taken place to minimise the risk for reverse causation. We applied HEIDI outlier analysis as part of the GSMR analysis to minimise the risk of horizontal pleiotropy. Comparison with five other popular summary-level MR methods showed broad concordance with our results. It should be noted that PBMC proteomics were measured on a smaller cohort than the plasma proteomics; subsequently in most cases only one SNP was utilised for the MR estimate. This may have led to a limited statistical power of detection in the PBMC MR results. However, previous work has suggested that the proportion of sample size needed for the genetic-variant-to-exposure GWAS can be less than that of the genetic-variant-to-outcome GWAS [26]. Finally, for both mass-spectrometry and affinity-based proteomics, interpretation of abundance measurements in the context of protein altering genetic variation is very challenging.

Recent technological and logistical advances have made population-level plasma proteome analysis possible, to this we have added the cellular proteome of PBMCs. Instrumental variable analyses, like MR, offer a practical alternative to randomised control trials for screening large numbers of proteins in the search for potential therapeutic protein targets; positioning, with careful interpretation of the results, proteome-wide analysis as a cornerstone of future drug-discovery efforts, helping to prioritise targets for further investigation.

## 4 Methods

### 4.1 Generation Scotland PBMC preparation

Generation Scotland: Scottish Family Health Study (GS:SFHS) is a population and family-based cohort, recruited from the Scottish population, between 2006 and 2011 [5]. Peripheral blood mononuclear cell samples (PBMCs) were available from Generation Scotland for 862 individuals. PBMC samples were available for a sample of those recruited in Glasgow or Dundee with parents born in Scotland, no further selection criteria were applied.

#### 4.1.1 Protein measurement

PBMCs were isolated from approximately 5ml whole blood that had been collected in acid-citrate-dextrose. Separation was performed using density gradient separation (Histopaque-1077; Sigma-Aldrich) by the European Collection of Authenticated Cell Cultures (ECACC) using a standardised protocol. PBMCs were then suspended in foetal-calf serum with 10% DMSO, frozen in a rate-controlled manner, and stored in liquid nitrogen until withdrawn for this study. Sample preparation and mass-spectrometry is described elsewhere [27], and summarised here for clarity.

Frozen PBMC cell pellets were retrieved from liquid nitrogen storage, thawed and washed in phosphate-buffered saline prior to lysis in 40*µ*L of 6M guanidine hydrochloride with 100mM tris(hydroxymethyl)aminomethane (‘lysis buffer’) and sonicated. 2*µ*l of sample was taken for a protein assay (Pierce BCA protein assay, Thermo-Fisher; Catalog number 23227). Protein concentration, per sample, was then standardised to a ceiling of 15*µ*g of protein in 20*µ*L lysis buffer. Samples were reduced and alkylated (1*µ*L of 100mM tris-carboxyethylphosphine, 1*µ*L of 200mM chloroacetamide, per sample) and heated for 5 minutes at 90-95*^◦^*C.

Proteins were digested with LysC (300ng per sample, overnight digest, 37*^◦^*C; Wako) and trypsin (150ng per sample, 4 hour digest, 37*^◦^*C; Pierce, Thermo Fisher). Digestion was stopped by the addition of 16*µ*L 10% trifluoroacetic acid (TFA), per sample.

Peptides were desalted on C18 stage tips. Stage tips were activated with 15*µ*L methanol, washed with 50*µ*L 0.1% TFA (pre- and post- sample loading), and eluted with 40*µ*L 80% acetonitrile + 0.1% TFA. Following elution, samples were dried and resuspended in 14*µ*L mass-spectrometry grade water. 9*µ*L was acidified with 1*µ*L 1% TFA and stored frozen prior to mass-spectrometry analysis.

Samples were prepared for mass-spectrometry in 20 batches (19 batches of 43 samples, and one batch of 45), and (where possible) one batch of sample repeats where the original preparation had failed (6 samples). Each batch included a pooled standard as well as within and between batch repeats.

#### 4.1.2 Mass-spectrometry

LC-MS/MS was performed on a Thermo Ultimate 3000 RSLC Nano UPLC coupled to a Thermo Fisher Q Exactive plus mass-spectrometer (Thermo Fisher). Samples were directly injected from a 96-well plate onto an Aurora UHPLC column from IonOpticks (Ion Opticks Pty Ltd). A Proxeon nano-spray ionisation source (Proxeon Biosystems) with a capillary temperature of 250°C and an optimised voltage of 1.4-1.7kV was used. A 120 minute gradient (2%-30% B in 110 min, 30%-45% B in the next 10 min; A=2% acetonitrile, B=80% acetonitrile, 0.5% acetic acid throughout; the composition was raised to 100% B in 7 minutes after the analytical gradient to wash the column, and total equilibration time was 20 minutes). Data-dependent acquisition (DDA), was run with a scan range of 350 to 1400 m/z using the Orbitrap at a resolution of 70,000 in profile mode. The top 24 parent peaks were selected for fragmentation. HCD fragmentation was performed with a normalised collision energy of 26 and spectra were acquired in centroid mode at a resolution of 17,500. Charge states accepted for MS2 were 2-5, peptide match was preferred and dynamic exclusion was 30 seconds. A single injection was performed for all batches, and a second injection on a separate occasion for 75% (15/20) of them.

#### 4.1.3 Data searching and annotation

Searches were conducted per batch. Data were processed using MaxQuant(v1.6.5.0) [28], matching against UniProt human (9606) reference proteome (5640) release 2019 11, canonical sequence and isoforms [29].

The search was performed for trypsin-digested peptides with up to two permitted missed cleavages. The fixed modification carbamidomethyl (C), and the variable modifications oxidation (M), acetylation (protein N-terminus), and methyl (KR;E) were included. Match-between-runs was allowed. MaxQuant built-in label-free quantification was not used. A false discovery rate of the peptide-spectrum matches of 0.01 was used for the search.

#### 4.1.4 Imputation

Missing data were imputed, per batch, using IceR (version 0.9.12) [30]. Parameter settings used can be found in the Supplementary Information 1: IceR Parameters. Once imputation was complete, quality control was performed. Batches with any of the following were removed: 1) an inconsistent gradient length (1 batch run) or low quality chromatography (1 batch run); 2) a median RT-deviation across the batch of *>* 1 min (4 batches runs); or 3) an FDR of *>* 5% of IceR quantification (when compared to 500 randomly chosen MaxQuant quantifications) (5 batch runs).

Twenty-four batch runs passed QC (17 unique batches: 10 with a single injection, 7 with two). A single injection of the repeats plate (6 samples) was included. In total, 1,068 mass-spectrometry runs, from 736 unique individuals, passed quality control.

#### 4.1.5 Normalisation

Intensity measurements were normalised using variance stabilisation normalisation in the R package ‘vsn’ (version 3.62.0) [31, 32]. The fit model (‘vsn2’) was computed using non-imputed intensities only (that is, prior to imputation with IceR) and applied (‘predict’) to both imputed and non-imputed data combined.

As a result of necessary sample preparation, ‘features’ measured during a mass-spectrometry proteomics experiment correspond to different charge states and isotopes of short post-translational modification-specific peptide sequences. The abundance of these features then need to be combined in order to quantify the abundance of the post-translational modification-specific peptides themselves, and the protein groups from which they could have originated.

Each mass-spectrometry feature was mapped to a post-translational modification-specific peptide sequence by MaxQuant. These features were combined with label-free quantification (LFQ; Section: ‘Extraction of Maximum Peptide Ratio Information’ [33]) and the resulting entity referred to as a **‘modification-specific peptide’** throughout this manuscript.

Each feature was further mapped by MaxQuant (‘All Peptides’ table) to a group of isoform-specific proteins from which is could have originated. As mass-spectrometry measures short peptide sequences, it is common for a peptide sequence to potentially have originated from multiple proteins; for example, it may be present in a shared protein domain. This means that each peptide maps to a group of proteins, rather than uniquely to a single one. Considering Swiss-Prot proteins only, the features mapped to each unique protein group were combined using LFQ, and the resulting grouping referred to as an **‘isoform-specific protein group’** throughout this manuscript.

#### 4.1.6 pQTL assessment (GWAS Model)

A mixed-linear model was performed using the GENESIS (v2.22) R package [34]. The following were included as fixed effects 1) age, 2) sex, 3) age squared, and 4) 20 genetic principal components, and as random effects: 1) genetic relationship matrix (GRM), 2) peptide digest batch, 3) mass-spectrometry batch, and 4) sample replicates. In this context, a ‘sample replicate’ is a sample that was processed from cell lysate multiple times, either within or between batches.

Genomic PCs and the GRM were created using the ‘pcair’ and ‘pcrelate’ functions from the GENESIS (v2.22) package and the ‘snpgdsIBDKING’ and ‘snpgdsLDprun-ing’ functions from the SNPRelate (v1.26) package [35]. Where the matrix denoting a random-effect was invariant it was removed from the model.

#### 4.1.7 Quality control: Genotyping

The HRC v1.1 imputation was used [36], as described for Generation Scotland [37]. SNPs that had an info score of *>* 0.9 and a minor-allele frequency *>* 0.01 in a random set of 861 of the individuals with PBMC samples available were considered in the GWAS: in total, 6,804,554 variants.

#### 4.1.8 Quality control: GWAS

Isoform-specific proteins/modification specific peptides that had been measured in 400, or more, unique individuals, and had a GWAS genomic inflation (lambda) of *<* 1.1 were considered for further analysis. In total, 5,114 isoform-specific protein groups (Supplementary Table 6), and 29,639 modification-specific peptide GWAS were considered. Where relevant, cis regions were identified (± 1Mb of the gene) for those isoform-specific protein groups that mapped to a single Swiss-Prot ID, and that that ID mapped to a single gene.

#### 4.1.9 Missense variation mapping

Measured peptide fragments were first mapped to their corresponding Ensembl transcript ID by using BLAT v.37x1 [38] against the set of all peptides translated from Ensembl genes (available at: https://ftp.ensembl.org/pub/grch37/release-108/fasta/homosapiens/pep/Homosapiens.GRCh37.pep.all.fa.gz; accessed: 26 October 2022). They were then mapped based on Ensembl transcript ID to their corresponding genomic coordinates, which were extracted from Ensembl using BioMart (https://grch37.ensembl.org/; accessed: 26 October 2022).

Annotated SNPs catalogued by dbSNP [39] (dbSNP build 155; available at: https://ftp.ncbi.nlm.nih.gov/snp/latest_release/VCF/GCF_000001405.25.gz; accessed: 26 October 2022) were filtered for missense variants using BCFtools v.1.13 [40]. Only SNPs with MAF above 0.05 in the 1,000 Genomes GBR subpopulation were retained [24]. Missense variants located within the mapped genomic regions were obtained using the ‘intersect’ command from BedTools v.2.30.0 [41]. Unmapped peptides and peptides mapped to multiple genomic locations were removed.

### 4.2 UK Biobank plasma proteins

The UK Biobank is a large-scale cohort study based in the United Kingdom, with a range of different measurements and data for approximately 500,000 individuals. The UK Biobank Pharma Proteomics Project (UKB-PPP) is a pre-competitive bio-pharmaceutical consortium formed in order to produce the plasma proteomic profiles of 54,219 individuals from the UKB. They identified 2,941 unique proteins in their data. Data preparation, quality control and GWAS summary creation can be found elsewhere [6]. GWAS summary files for all 2,941 proteins, on the European (discovery) cohort, were downloaded from the Synapse data storage platform [6, 42]. These files were used in the GSMR procedure.

As 6 proteins (CXCL8, IDO1, IL6, LMOD1, SCRIB and TNF) were measured on multiple protein panels, we only retained the cardiometabolic panels to ensure no proteins were duplicated, with the exception of SCRIB for which no cardiometabolic panel existed – the oncology panel was retained. This filtering gave 2,923 proteins. Following this we only retained cis-genetic-variants, those within ± 1Mb from the gene encoding region, for each protein which was significant below a Bonferroni corrected genome wide significance threshold (*p <* 5 × 10*^−^*^8^*/*2923). This left 1,913 proteins which had at least one instrument. Additionally 3 proteins were removed due to inability to map between GRCh38 and GRCh37 (PRSS2, PECAM1 and KIR2DL2). NTproBNP was removed due to its complexity when attempting to map. We considered proteins mapped to a single gene on an autosome only.

Some proteins measured in the UK Biobank proteomics dataset (AMY1A, BOLA2, CGB3, CKMT1A, DEFA1, DEFB104A, EBI3, FUT3, LGALS7, and MICB) originate from multiplexed analytes that capture signal from multiple genes using a single assay. In line with the UKB-PPP annotation, these assays are treated as one measurement, and we retained only the gene symbols listed above for consistency in naming and mapping. It was not possible for these entries to be decomposed further for gene-specific interpretation.

This resulted in 1,909 proteins remaining for GSMR and colocalisation analyses.

### 4.3 LD reference

An unrelated set of individuals was selected from Generation Scotland [37] for use as an LD reference and reporting allele frequencies. The ‘–make-king-table’ flag in PLINK was used to determine the relatedness of each of the samples. For sample pairs with a relatedness score of 0.025, or greater, one of the samples was removed to leave an unrelated subset of 6,862 samples. SNPs reported as significant and used in subsequent analyses from the PBMC GWAS were filtered to those with a minor allele frequency *>* 0.05 within this unrelated set of individuals. This LD reference was used for the GSMR runs with both the PBMC and plasma data.

### 4.4 Outcome GWAS process

GWAS were run on the following phenotypes from UKB: angina, type II diabetes, heart attack, high cholesterol and essential hypertension. A non-overlapping subset of UKB data was used to conduct the GWAS, such that no individual with proteomics data was used – ensuring minimal inflation from inclusion of those used in the plasma proteomics GWAS. Additionally filtering was used to select only those who were white British. Those who recorded an ICD-10 code before their date of assessment centre (when the blood sampling was conducted) or who self-reported at baseline the equivalent condition under self-report field 20002 were excluded, in order to conserve temporality.

Details of final cases and controls, along with field IDs for UKB data fields are provided in Table 3. Genome-wide association analyses were performed using REGENIE (v3.2.2), which implements a two-step approach for efficient mixed model analysis. In Step 1, the model was fit using directly genotyped autosomal genetic-variants, with a block size of 80,000 genetic-variants and a binary trait model. In line with similar GWAS studies [43], the model included as fixed effects: (1) age, (2) sex, (3) age squared, (4) assessment centre, (5) genotyping array, and (6) the top 20 genetic principal components. Population structure and relatedness were accounted for in REGENIE’s first step using ridge regression on common variants, employing a leave-one-chromosome-out (LOCO) framework.

**Table 3:** Table presenting the phenotypes used in this study. Each row provided the phenotype name, UKB data codes and the number of controls and cases used for each GWAS.

| Outcome | UKB Coding | No. of Controls | No. of Cases |
| --- | --- | --- | --- |
| Angina | I20 | 356,330 | 12,894 |
| Type II Diabetes | E11 | 353,474 | 22,492 |
| Heart Attack | I21 | 366,947 | 8,458 |
| High Cholesterol | E780 | 297,037 | 28,973 |
| Essential Hypertension | I10 | 262,964 | 89,998 |

In Step 2, imputed BGEN files, with a minimum information score of 0.9, and a minor allele count filter of 10 were used to test genetic-variant-trait associations using linear regression on the Step 1 predictions, again with the same covariates. All participants were of white British ancestry and passed UK Biobank’s standard QC filters.

### 4.5 GSMR

GSMR results were generated using GCTA 1.94 [9, 44]. The pQTLs (PBMC or plasma) were used as the exposure, and each of the selected UK Biobank CVD, and CVD-related risk-factors as the outcome. The unrelated set of individuals from Generation Scotland was used as the LD reference.

In the GCTA software three items are specified, namely (i) direction of GSMR, we specify ‘0’ for a forward direction assuming the exposure is directed to the outcome; (ii) maximum difference in allele frequency of each genetic-variant between the data inputs, which we set to ‘1.0’ to allow for any difference between the GWAS and the LD reference sample; (iii) minimum number of genome-wide significant and independent genetic-variant, set to ‘1’ in our case to allow for any number of genetic-variants to be used, recognising this is below the rule of thumb of 10 suggested by the GSMR authors [9]. Explicitly, the following settings were used in GCTA: –gsmr-direction 0 –diff-freq 1.0 –gsmr-snp-min 1.

Via built-in settings of the GCTA GSMR software, genetic instruments were selected based on genome-wide significance (*p <* 5 × 10*^−^*^8^), however please note our pre-filtering on p-value (Fig. 2), with linkage disequilibrium (LD) pruning conducted using the GSMR package. Instruments were clumped using an LD *r*^2^ threshold of 0.05 to ensure independence. These criteria aimed to minimise bias due to LD structure and ensure consistency across datasets.

The built-in HEIDI (Heterogeneity in Dependent Instruments) outlier procedure was run prior alongside the GSMR analysis to identify and remove potentially pleiotropic genetic-variants. HEIDI tests whether the genetic-variant-specific effect estimates are consistent across instruments by evaluating the heterogeneity of the genetic-variant-outcome associations conditional on their exposure associations. Under the assumption of no horizontal pleiotropy, all valid instruments should estimate the same causal effect. Genetic-variants showing significant heterogeneity are considered outliers, indicative of horizontal pleiotropy. We used the programme-default p-value threshold of 0.01 for identifying outliers, as recommended by original authors [9]. To account for multiple testing across instruments, we applied the Benjamini–Hochberg procedure to control the false discovery rate at 0.05 [22].

We note 8 and 285 proteins were removed from PBMCs and plasma, respectively, as they had 0, 2, or 4 index genetic-variants following clumping and GSMR analysis was not completed due to limitations of the GSMR software.

### 4.6 Colocalisation analysis

‘Coloc’ version 5.2.2 [11], jointly tested five hypothesis under a Bayesian framework:

- *H*_0_: No association with either the exposure or outcome.
- *H*_1_: Association with only the exposure, but not the outcome.
- *H*_2_: Association with only the outcome, but not the exposure.
- *H*_3_: Association with both exposure and outcome, through two independent genetic-variants.
- *H*_4_: Association with both the exposure and outcome, through one shared genetic-variant.

Default setting were used within the ‘coloc’ R package provided by the author for the setting of the prior probabilities. Cis-only SNPs, those in ± 1Mb of the gene encoding region, were utilised.

### 4.7 Enrichment Analyses

Enrichment analyses were performed using StringDB [23]. For each of the outcome traits, the protein names of all GSMR-significant (FDR *<* 0.05) non-MHC (chr6:28477797-chr6:33448354 [45]) plasma proteins were input into StringDB, with the organism selected as ‘Homo sapiens’. The standard protein mapping performed by StringDB, between the input set and its database, was manually checked and, as no edits were required, accepted.

Enrichment analyses were conducted for plasma protein data only as the number of significant protein-disease links identified in the PBMC dataset was too small to yield robust, interpretable enrichment results. Further, given that isoform-specific protein groups were measured, the creation of an appropriate background on StringDB was not possible.

For the UKB plasma enrichment analysis, a bespoke background of all 1,624 post-filtering instrumented proteins from the plasma data was used. The final networks of proteins identified by StringDB were of sizes 10, 24, 4, 18, 52 for the traits angina, type II diabetes, heart attack, high cholesterol and essential hypertension respectively.

### 4.8 Sensitivity Analyses

To assess the robustness of the gene-outcome associations identified via GSMR, we conducted MR analyses with five alternative, commonly used, MR methods: inverse-variance weighted (IVW) [46], MR-Egger regression [47], weighted median [48], weighted mode [49], and MR-RAPS [50]. These methods were applied to both the PBMC and plasma datasets independently.

Full results were tabulated in Supplementary Table 7 for the PBMC dataset and Supplementary Table 8 for the plasma dataset. To visualise the consistency across methods, we construct heatmaps (Supplementary Figures 6 and 7) indicating whether the direction of effect was concordant or discordant with GSMR, and whether the association was statistically significant (FDR *<* 0.05) in each method, for each gene-outcome pair identified as significant (FDR *<* 0.05) via GSMR for the PBMC data.

Each cell in the heatmaps represents a single method–gene–outcome combination and is colour-coded to denote concordance (purple) or discordance (yellow) with the GSMR estimate. The significance status is annotated with symbols: a filled circle for concordant and significant, a filled square for concordant and not significant, a filled cross for discordant and significant, and a light cross for discordant and not significant. This approach provides a visual summary of consistency across MR methods, highlighting findings that are robust to methodological variation.

To assess the robustness of our Mendelian randomisation (MR) findings to differences in case prevalence across cohorts, we performed an empirical sensitivity analysis using random down-sampling. For each trait, we generated modified genome-wide association study (GWAS) datasets by randomly down-sampling cases from the UK Biobank exposure and outcome cohorts to match the prevalence observed in the Generation Scotland PBMC proteomics cohort. For each resampled dataset, we re-ran both the SNP–exposure and SNP–outcome GWAS analyses, the latter using REGENIE with Firth correction to account for potential bias arising from unbalanced case–control ratios [51–53]. The resulting summary statistics were then used to repeat the GSMR analyses, allowing us to confirm that the observed causal estimates were not affected by the observed differences in disease prevalence between cohorts (Supplementary Figures 8 and 9).

## 5 Declarations

### 5.1 Ethics

Generation Scotland participants provided written informed consent. Generation Scotland was granted Research Tissue Bank status by the East of Scotland Research Ethics Service committee (REC reference 15/ES/0040, 25/ES/0013). This project was approved by Generation Scotland as reference GS18318. The data / material transfer agreement was made on 05 Feb 2019. Researchers on this project had access to genotype data and pseudo-anonymised research data and materials.

UK Biobank has approval from the North West Multi-centre Research Ethics Committee (REC reference 21/NW/0157; renewal of the original REC reference 11/NW/0382). Before being enrolled in the UK Biobank study, all participants provided written consent after being fully informed, adhering to the principles outlined in the Declaration of Helsinki. This research has been conducted using the UK Biobank Resource under Application Number 91924. The data and material transfer agreement was active from 21 Dec 2022. Researchers on this project had access to genotype data and pseudo-anonymised medical and biological data.

### 5.2 Data Availability

Summary statistics for the significant (*p <* 5 × 10*^−^*^8^*/*5114; Bonferroni correction) results from the isoform-specific Swiss-Prot protein group GWAS are available in Supplementary Table 9, and those with a p-value *<* 5 × 10*^−^*^8^ in the cis region (± 1Mb) of the gene to which protein group was mapped are available in Supplementary Table 10.

Full GWAS summary statistics of the isoform-specific Swiss-Prot protein groups and modification-specific peptides are available via Generation Scotland, and individual level data are available to bona fide researchers, subject to approval by the Generation Scotland data access committee: https://genscot.ed.ac.uk/for-researchers/access.

This research has been conducted using the UK Biobank Resource under Application Number 91924. Details of UK Biobank’s data access procedures are available here: https://www.ukbiobank.ac.uk/use-our-data/apply-for-access/.

### 5.3 Author Contributions

**CAO**: Data curation; Formal analysis; Investigation; Software; Visualization; Writing – original draft; Writing – review & editing. **JN**: Formal analysis; Investigation; Writing – original draft; Writing – review & editing. **JW**: Conceptualization; Data curation; Formal analysis; Investigation; Methodology; Project administration; Resources; Software; Validation; Writing – original draft; Writing – review & editing. **AR**: Data curation; Formal analysis; Investigation; Software; Writing – review & editing. **TR**: Investigation; Writing – review & editing. **SCH**: Investigation; Writing – review & editing. **AC**: Conceptualization; Data curation; Resources; Writing – review & editing. **JKB**: Conceptualization; Project administration; Resources; Supervision; Writing – review & editing. **AvK**: Conceptualization; Data curation; Investigation; Methodology; Project administration; Resources; Supervision; Writing – review & editing. **CH**: Conceptualization; Funding acquisition; Methodology; Project administration; Resources; Supervision; Writing – review & editing. **AK**: Conceptualization; Formal analysis; Investigation; Methodology; Software; Supervision; Validation; Writing – original draft; Writing – review & editing. **SVB**: Conceptualization; Data curation; Formal analysis; Funding acquisition; Investigation; Methodology; Project administration; Software; Supervision; Validation; Writing – original draft; Writing – review & editing. **ADB**: Conceptualization; Data curation; Formal analysis; Funding acquisition; Investigation; Methodology; Project administration; Supervision; Validation; Writing – original draft; Writing – review & editing.

## Acknowledgements

This study would not have been possible without the participants and staff of both Generation Scotland and UK Biobank, for which we are very grateful.

## 5.4 Funding

CAO was supported by the EPSRC Centre for Doctoral Training in Mathematical Modelling, Analysis and Computation (MAC-MIGS) funded by the UK Engineering and Physical Sciences Research Council (grant EP/S023291/1), Heriot-Watt University and The University of Edinburgh.

TR would like to acknowledge strategic funding grant BBSRC Institute Strategic Programme grants (BBS/E/D/20002172 and BBS/E/D/20002174).

SCH would like to acknowledge the BBSRC Strategic Programme Grant to the Roslin Institute (BB/P013732/1, BB/P013759/1).

JKB gratefully acknowledges funding support from a Wellcome Trust Senior Research Fellowship (223164/Z/21/Z), UKRI grants (MR/Y030877/1, MC PC 20004, MC PC 19025, MC PC 1905, MRNO2995X/1, and MC PC 20029), Sepsis Research

(Fiona Elizabeth Agnew Trust), a BBSRC Institute Strategic Programme Grant to the Roslin Institute (BB/P013732/1, BB/P013759/1) and support of Baillie Gifford and the Baillie Gifford Science Pandemic Hub at the University of Edinburgh.

AK is supported by a Langmuir Talent Development Fellowship from the Institute of Genetics and Cancer, and a philanthropic donation from Hugh and Josseline Langmuir.

AK and SVB acknowledge support of the UKRI AI programme, and the Engineering and Physical Sciences Research Council, for CHAI - EPSRC AI hub for Causality in Healthcare AI with real data [grant number EP/Y028856/1].

ADB would like to acknowledge funding from the Wellcome PhD training fellowship for clinicians (204979/Z/16/Z), the Edinburgh Clinical Academic Track (ECAT) programme, and UKRI grant MR/Y030877/1.

CH and AR would like to acknowledge the MRC University Unit Programme Grant to the Human Genetics Unit (MC UU 00007/10).

Generation Scotland received core support from the Chief Scientist Office of the Scottish Government Health Directorates [CZD/16/6] and the Scottish Funding Council [HR03006]. Genotyping of the GS:SFHS samples was carried out by the Genetics Core Laboratory at the Wellcome Trust Clinical Research Facility, Edinburgh, Scotland and was funded by the Medical Research Council UK and the Wellcome Trust (Wellcome Trust Strategic Award “STratifying Resilience and Depression Longitudinally” (STRADL) Reference 104036/Z/14/Z).

This work has made use of the resources provided by the Edinburgh Compute and Data Facility (ECDF) (http://www.ecdf.ed.ac.uk/) with support from MRC grant MC PC MR/X013677/1.

The funders had no role in study design, data collection and analysis, decision to publish, or preparation of the manuscript.

For the purpose of open access, the authors have applied a CC BY public copyright licence to any Author Accepted Manuscript version arising from this submission.

## 5.5 Disclosure and competing interest statement

The authors declare that they have no conflict of interest.

## 6 Supplementary Materials

### Supplementary Table 1

**Table 1:** Demographics and Cardiovascular Disease Prevalence/Incidence.

| Cardiovascular Condition<br>(ICD-10/ICD-9 Code) | Prevalence<br>Generation<br>Scotland<br>Proteomics,<br>N (%) | Prevalence<br>UK Biobank<br>Proteomics,<br>N (%) | Incidence<br>UK Biobank<br>Outcome<br>N (%) |
| --- | --- | --- | --- |
| Angina (I20/413) | 15 (1.74%) | 3,407 (7.36%) | 12,894 (3.49%) |
| Type II Diabetes (E11/250.00) | < 10 (< 1.16%) | 3,954 (8.54%) | 22,492 (5.98%) |
| Heart Attack (I21/410) | < 10 (< 1.16%) | 1,776 (3.83%) | 8,458 (2.25%) |
| High Cholesterol (E780/272.0) | < 10 (< 1.16%) | 7,357 (15.88%) | 28,973 (8.89%) |
| Essential Hypertension (I10/401) | 30 (3.48%) | 15,507 (33.48%) | 89,998 (25.50%) |
| Population Demographics |  |  |  |
| Age, Mean (SD) | 52 (9.04) | 57 (8.10) | 57 (7.99) |
| Sex Ratio, N Male (%) | 347 (40.26%) | 24,928 (53.82%) | 215,261<br>(54.34%) |

This table provides descriptive statistics for the individuals included in the proteomics analyses from Generation Scotland (GS) and UK Biobank (UKB), and those individuals in UKB used in the outcome GWA, alongside rates for the five cardiovascular or cardiovascular-related conditions investigated using GSMR in each set. **Generation Scotland (GS):** Values are for the 862 participants with peripheral blood mononuclear cell (PBMC) samples available in Generation Scotland, and included in this study. Disease prevalence is recorded at the time of blood sampling. **UK Biobank (UKB) left:**Disease prevalence in the UKB proteomics cohort recorded at the point of blood sample extraction. **UK Biobank right:** Values are incident cases (between enrolment and analysis) for participants used in the outcome GWA analyses. Specifically, case numbers reflect those used in the outcome GWAS for GSMR, where the phenotype occurred after protein sampling, ensuring temporal ordering between exposure and outcome. Demographics (age at recruitment and sex) reflect the white British, non-proteomics sub-cohort on which these outcome GWAS were run.

#### Supplementary Table 2

**GS PBMC GSMR and colocalisation results.** The full results for all PBMC measured proteins, for all five CVD/CVD-risk related outcomes. The included columns are: ‘trait’, ‘Gene name’, ‘Swiss-Prot ID’, ‘Gene chromosome’, ‘Gene start (bp)’, ‘bxy’, ‘p’, ‘-log(p)’, ‘accept’, ‘nsnp’, ‘H0’, ‘H1’, ‘H2’, ‘H3’ and ‘H4’. The trait refers to the outcome, whilst the gene name refers to the protein tested along with its corresponding known chromosome and starting location. ‘Swiss-Prot ID’ is the Swiss Prot IDs present in the isoform-specific protein group. ‘bxy’ represents the effect size determined by the GSMR procedure, ‘p’ represents the p-value of the GSMR procedure and ‘accept’ determines whether the protein is accepted at a Benjamini–Hochberg FDR rate of 0.05, and ‘nsnp’ refers to the number of instruments used in the GSMR. Finally H0-H4 relate to the posterior probabilities of the colocalisation tests.

#### Supplementary Table 3

**UKB plasma GSMR and colocalisation results.** The full results for all plasma measured proteins, for all five CVD/CVD-risk related outcomes. The included columns are: ‘trait’, ‘Gene name’, ‘Gene chromosome’, ‘Gene start (bp)’, ‘bxy’, ‘p’, ‘-log(p)’, ‘accept’, ‘nsnp’, ‘H0’, ‘H1’, ‘H2’, ‘H3’ and ‘H4’. The trait refers to the outcome, whilst the gene name refers to the protein tested along with its corresponding known chromosome and starting location. ‘bxy’ represents the effect size determined by the GSMR procedure, ‘p’ represents the p-value of the GSMR procedure and accept determines whether the protein is accepted at a Benjamini–Hochberg FDR rate of 0.05, and ‘nsnp’ refers to the number of instruments used in the GSMR. Finally H0-H4 relate to the posterior probabilities of the colocalisation tests.

#### Supplementary Table 4

**UKB plasma enrichment analyses.** Full enrichment analyses using StringDB, provided as one table across all traits. The background for all of these networks were the 1,624 proteins instrumented in the plasma data set. No enrichment pathways were found for the Diabetes and Hypertension traits.

#### Supplementary Table 5

COMT Locus Zoom and Lead SNP Information

**Table 5:** Lead SNP for COMT found to be significant in both the PBMC and plasma GSMR results. . The SNP shown, is the one selected as the lead SNP/only-instrument in the PBMC GSMR (smallest p-value). It is observed that the direction of effect, as measured by the respective GWAS, on the protein was the same in both the PBMC and plasma data.

| Field | Value |
| --- | --- |
| Gene | COMT |
| Chromosome (Chr) | 22 |
| Position (Pos) | 19948863 |
| Effect Allele | C |
| Other Allele | G |
| $\beta_{\text{GS}}$ | -0.273 |
| $\text{SE}_{\text{GS}}$ | $2.500 \times 10^{-2}$ |
| $\text{p-value}_{\text{GS}}$ | $6.700 \times 10^{-28}$ |
| $\beta_{\text{UKB}}$ | -0.445 |
| $\text{SE}_{\text{UKB}}$ | $7.643 \times 10^{-3}$ |
| $\text{p-value}_{\text{UKB}}$ | $< 1 \times 10^{-30}$ |

#### Supplementary Table 6

**Isoform-Specific Protein Groups Tested in GWAS.** List of all 5,114 isoform-specific Swiss-Prot protein groups upon which GWA were considered. Tab-separated file. Columns: Proteins allpep spIso: isoform-specific Swiss-Prot protein group; N: Number of unique individuals present in the GWA.

#### Supplementary Table 7

**PBMC MR results across five alternative methods.** The full MR results for all PBMC-measured proteins, for all five CVD/CVD-risk related outcomes. The GSMR result are also provided for completeness. The included columns are: Gene name, Outcome, GSMR Effect, GSMR P, IVW Effect, IVW P, Egger Effect, Egger P, Weighted Median Effect, Weighted Median P, Weighted Mode Effect, Weighted Mode P, RAPS Effect, and RAPS P. Each column provides the effect size and corresponding p-value for a different MR method applied to the same gene–outcome pair. Note that the p-values obtained by MR-RAPS are set to zero if they are below the threshold of approximately 2.2 × 10*^−^*^16^; the p-values of the other methods are set to zero if they are below the threshold of approximately 10*^−^*^310^.

#### Supplementary Table 8

**Plasma MR results across five alternative methods.** Akin to the PBMC results, this tables gives full MR results for all plasma-measured proteins, for all five CVD/CVD-risk related outcomes. The GSMR result are, once again, provided for completeness. The included columns are: Gene name, Outcome, GSMR Effect, GSMR P, IVW Effect, IVW P, Egger Effect, Egger P, Weighted Median Effect, Weighted Median P, Weighted Mode Effect, Weighted Mode P, RAPS Effect, and RAPS P. Each column provides the effect size and corresponding p-value for a different MR method applied to the same gene–outcome pair. Note that the p-values obtained by MR-RAPS are set to zero if they are below the threshold of approximately 2.2 × 10*^−^*^16^; the p-values of the other methods are set to zero if they are below the threshold of approximately 10*^−^*^310^.

#### Supplementary Table 9

**Genome-wide PBMC pQTL results.** Supplementary Table 9 contains significant (GWA p-value *<* 5 × 10*^−^*^8^*/*5114; Bonferroni correction) results for isoform-specific, Swiss-Prot only, protein groups measured in ≥ 400 unique individuals with a GWA genomic inflation (lambda) *<* 1.1. SNPs are limited to those with a minor allele frequency *>* 0.05 in a set of unrelated individuals from Generation Scotland (relatedness score of *<* 0.025 for all pairwise comparisons).

Included columns are as follows: Proteins allpep spIso: Isoform-specific Swiss-Prot IDs in the group; N: Number of unique individuals included in the GWA study; lambda: Genomic inflation (lambda) of the GWA study; chr: Chromosome (GRCh37); pos: Position (GRCh37); Score: The value of the score function (as per the GENESIS v2.22 ‘assocTestSingle’ function); Score.SE: The estimated standard error of the Score (as per the GENESIS v2.22 ‘assocTestSingle’ function); Score.pval: The score p-value (as per the GENESIS v2.22 ‘assocTestSingle’ function); Est: An approximation of the effect-size estimate for each additional copy of the effect allele (as per the GENESIS v2.22 ‘assocTestSingle’ function); Est.SE: An approximation of the standard error of the effect-size estimate (as per the GENESIS v2.22 ‘assocTestSingle’ function); PVE: An approximation of the proportion of phenotype variance explained (as per the GENESIS v2.22 ‘assocTestSingle’ function); other.allele: Other allele in the GWA study; effect.allele: Effect allele in the GWA study; and effect.allele.frequency: The frequency of the effect allele from a set of unrelated individuals from Generation Scotland (relatedness score of *<* 0.025 for all pairwise comparisons).

#### Supplementary Table 10

**Cis-region PBMC pQTL results.** Considering only SNPs located within the cis region (±1Mb) of the gene to which protein group was mapped, Supplementary Table 10 contains significant (GWA p-value *<* 5 × 10*^−^*^8^) results for isoform-specific, Swiss-Prot only, protein groups measured in ≥ 400 unique individuals with a GWA genomic inflation (lambda) *<* 1.1. SNPs are limited to those with a minor allele frequency *>* 0.05 in a set of unrelated individuals from Generation Scotland (relatedness score of *<* 0.025 for all pairwise comparisions). The results are limited to those protein groups that mapped to a single UniProtKB ID (Swiss-Prot) and that that UniProt ID mapped to a single gene. Mapping was performed using https://grch37.ensembl.org/biomart/martview/ (accessed 25 May 2023).

The columns are as per Supplementary Table 9 with the following additions: Gene stable ID: Ensembl Gene ID of the mapped Gene; Gene start (bp): Gene start location; Gene end (bp): Gene end location; and Gene name: Gene name of the mapped Gene.

#### Supplementary Figures 1-4

**UKB plasma GSMR networks.** Supplementary Figures 1, 2, 3, and 4 show the String-DB network graphs for type II diabetes, heart attack, high cholesterol, and essential hypertension respectively, of all non-MHC region proteins identified by GSMR to have a significant relationship with the corresponding outcome, following a multiple testing correction (FDR *<* 0.05, Benjamini–Hochberg). Results on each network are shaded when the H4 colocalisation posterior probability, the hypothesis that both the exposure and outcome share a common associated genetic-variant, is above 0.5. The posterior probability for H4 is coloured when between 0.5 and 0.8 (blue) or larger than 0.8 (orange). A greater number indicates greater support for the hypothesis. Lines between nodes represent different forms of interaction evidence through: co-occurrence, co-expression, databases, experiments, gene fusion, neighbourhood and text-mining.

**Supplementary Figure 1:**
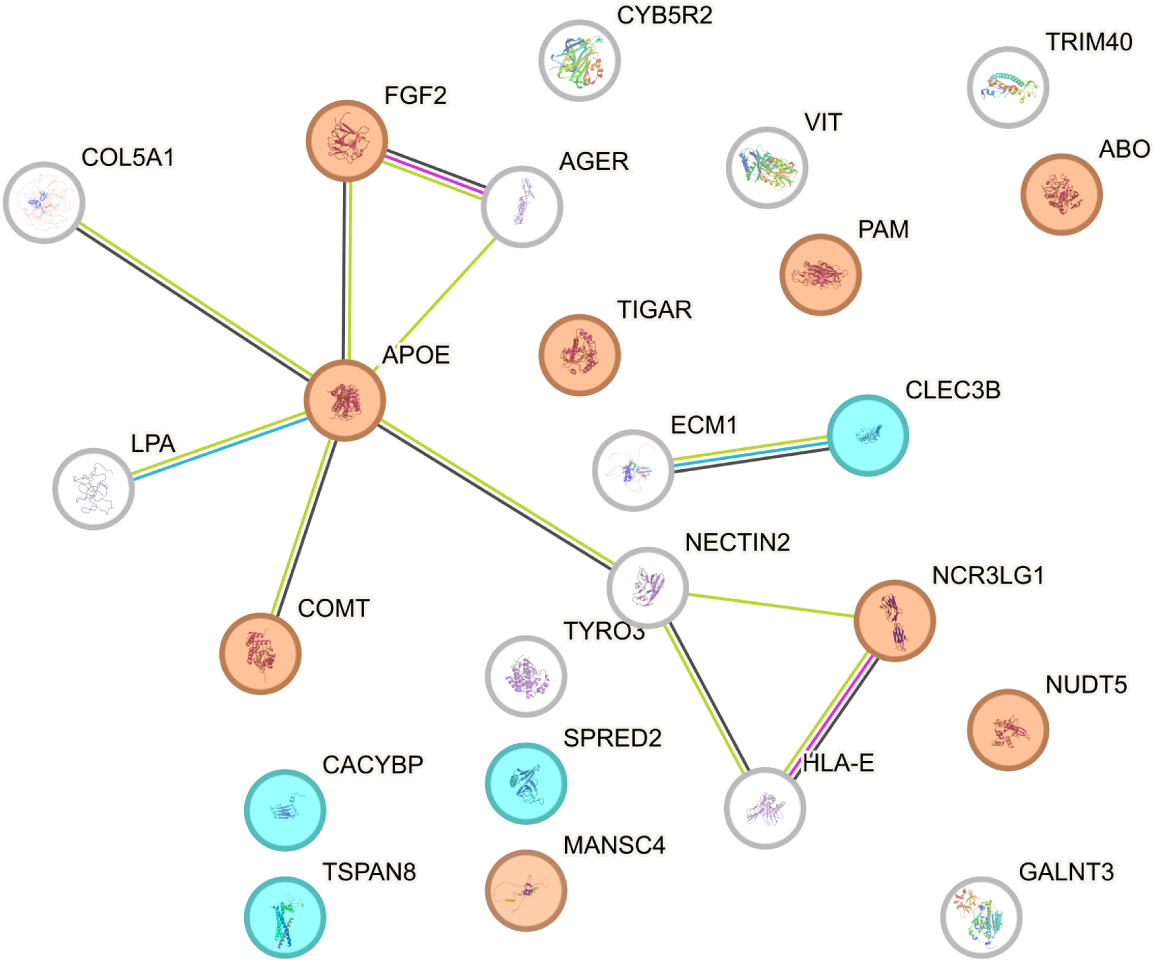
UKB GSMR and colocalisation results network for the **type II diabetes** outcome from StringDB.

**Supplementary Figure 2:**
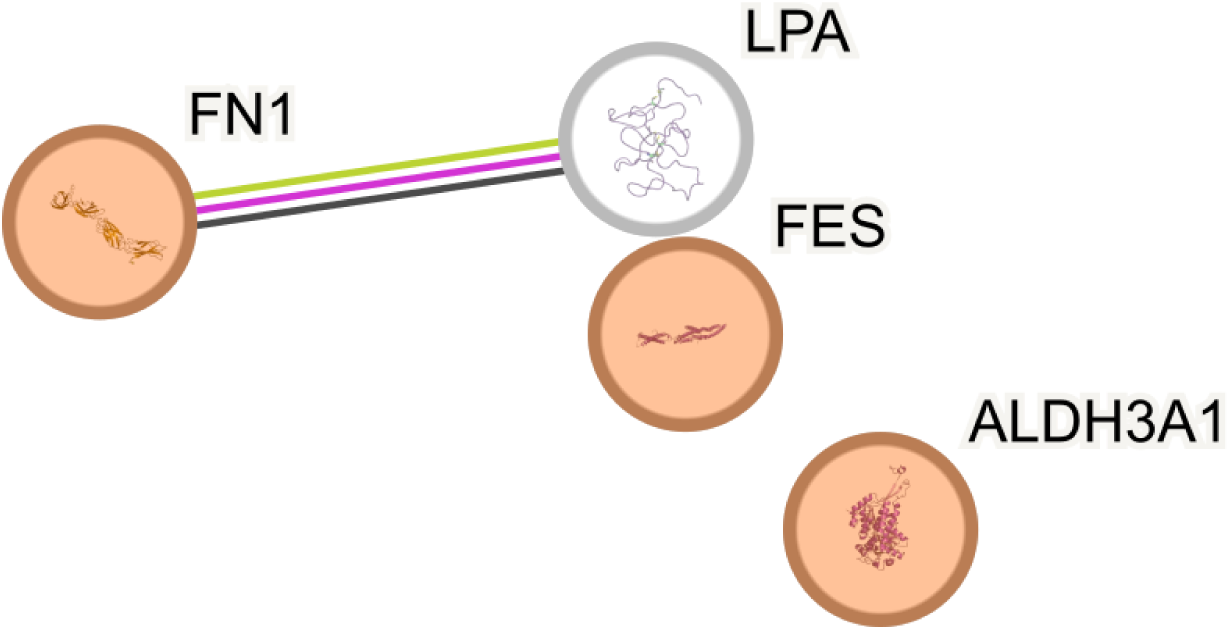
UKB GSMR and colocalisation results network for the **heart attack** outcome from StringDB.

**Supplementary Figure 3:**
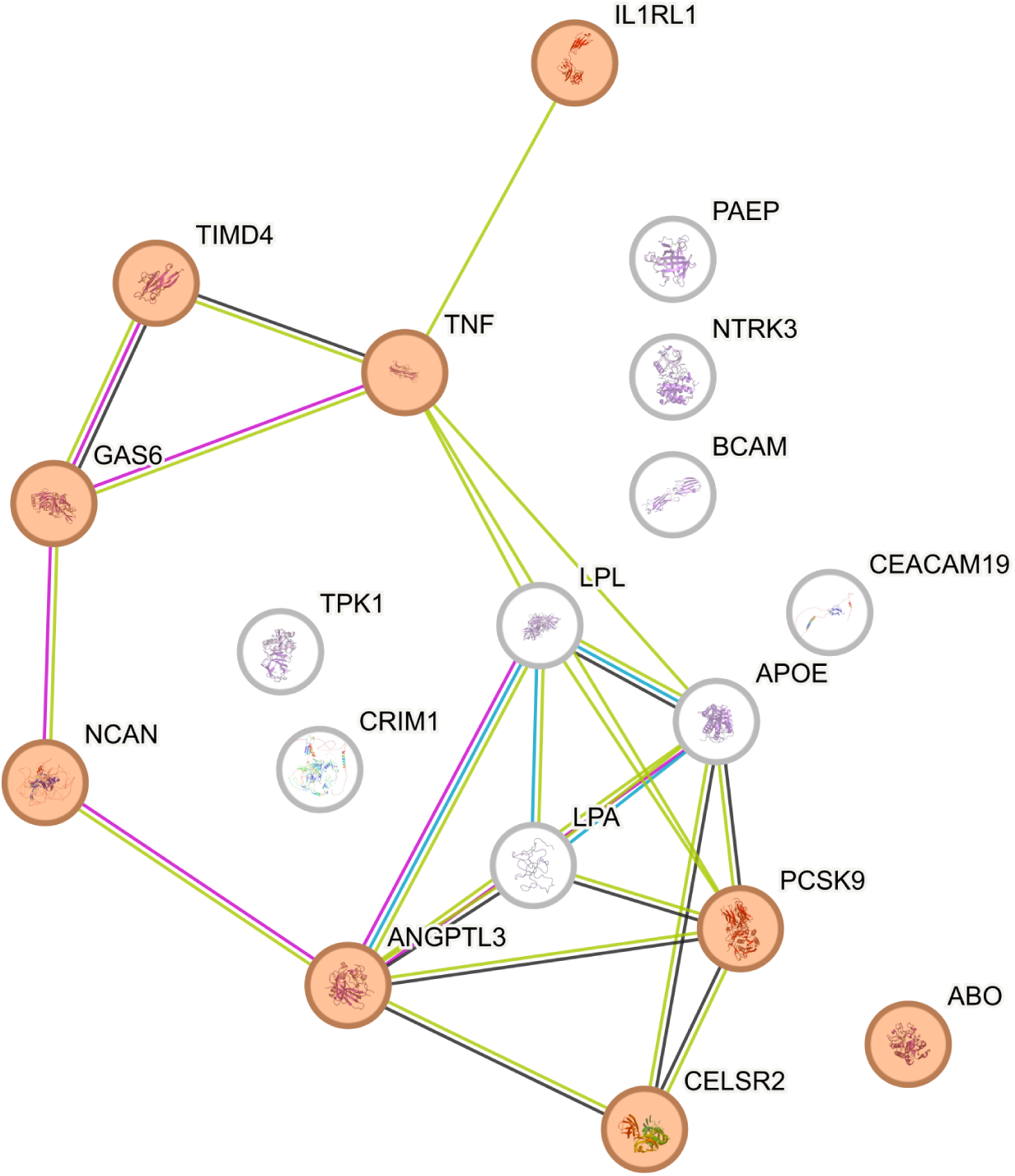
UKB GSMR and colocalisation results network for the **high cholesterol** outcome from StringDB.

**Supplementary Figure 4:**
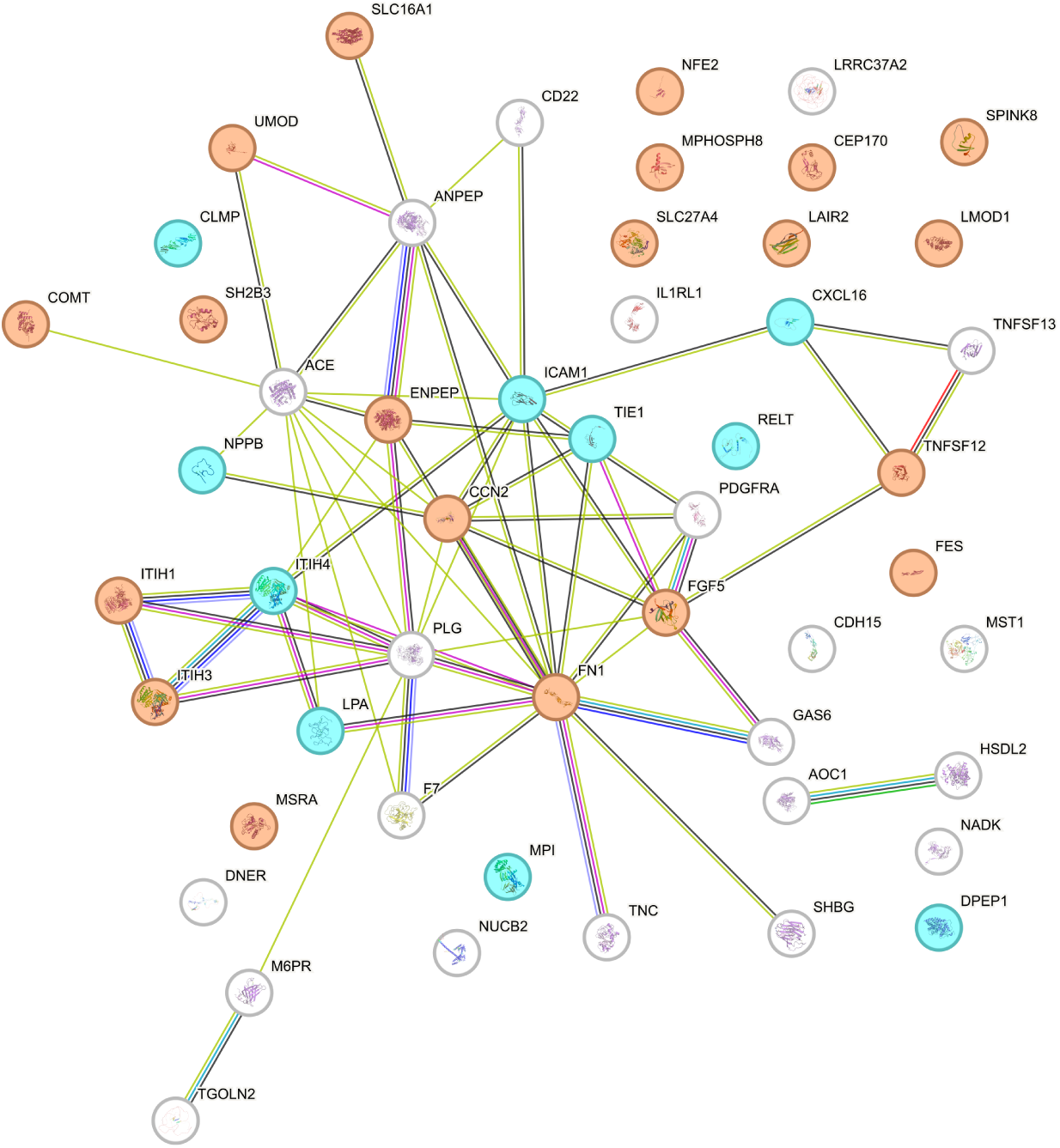
UKB GSMR and colocalisation results network for the **essential hypertension** outcome from StringDB.

#### Supplementary Figure 5

**Supplementary Figure 5:**
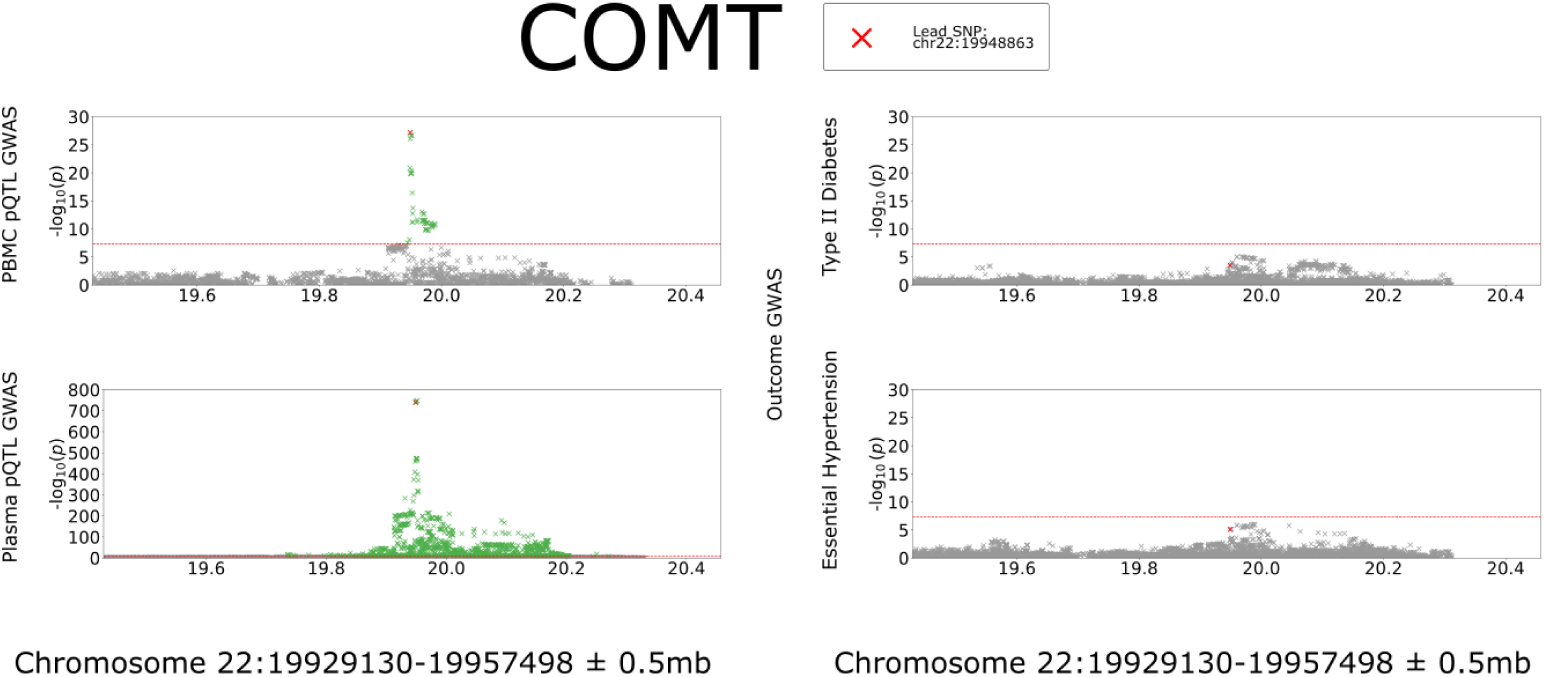
Locus Zoom of COMT for PBMC and plasma data, with outcomes where it was found to be significant in both PBMCs and Plasma. We plot the gene encoding it ±0.5Mb. Column 1, top row: peripheral-blood mononuclear cell pQTL results; column 1, bottom row: plasma pQTL results; column 2, top row: type II diabetes GWAS results; column 2, bottom row: essential hypertension GWAS results. The most significant SNP in the peripheral-blood mononuclear cell protein GWAS is marked as a red cross. The red line is at genome-wide significance (p = 5 × 10*^−^*^8^).

#### Supplementary Figures 6-7

In order to provide robust validation to our results presented here we perform sensitivity checks on the method. We run alternative MR methods: IVW, MR Egger, Weighted Median, Weighted Mode and MR RAPS, all of which use summary-level statistics from two samples to estimate the effect between an exposure and outcome in the MR framework. We do this to check concordance with direction of effect and significance with the results from the GSMR process. Figures 6 and 7 show the results for the proteins which were found significant in the GSMR processes for the PBMC and plasma proteomics (respectively), in the form of a heat-map. Full results from alternative MR methods can be found in Supplementary Table 8 and 9.

**Supplementary Figure 6:**
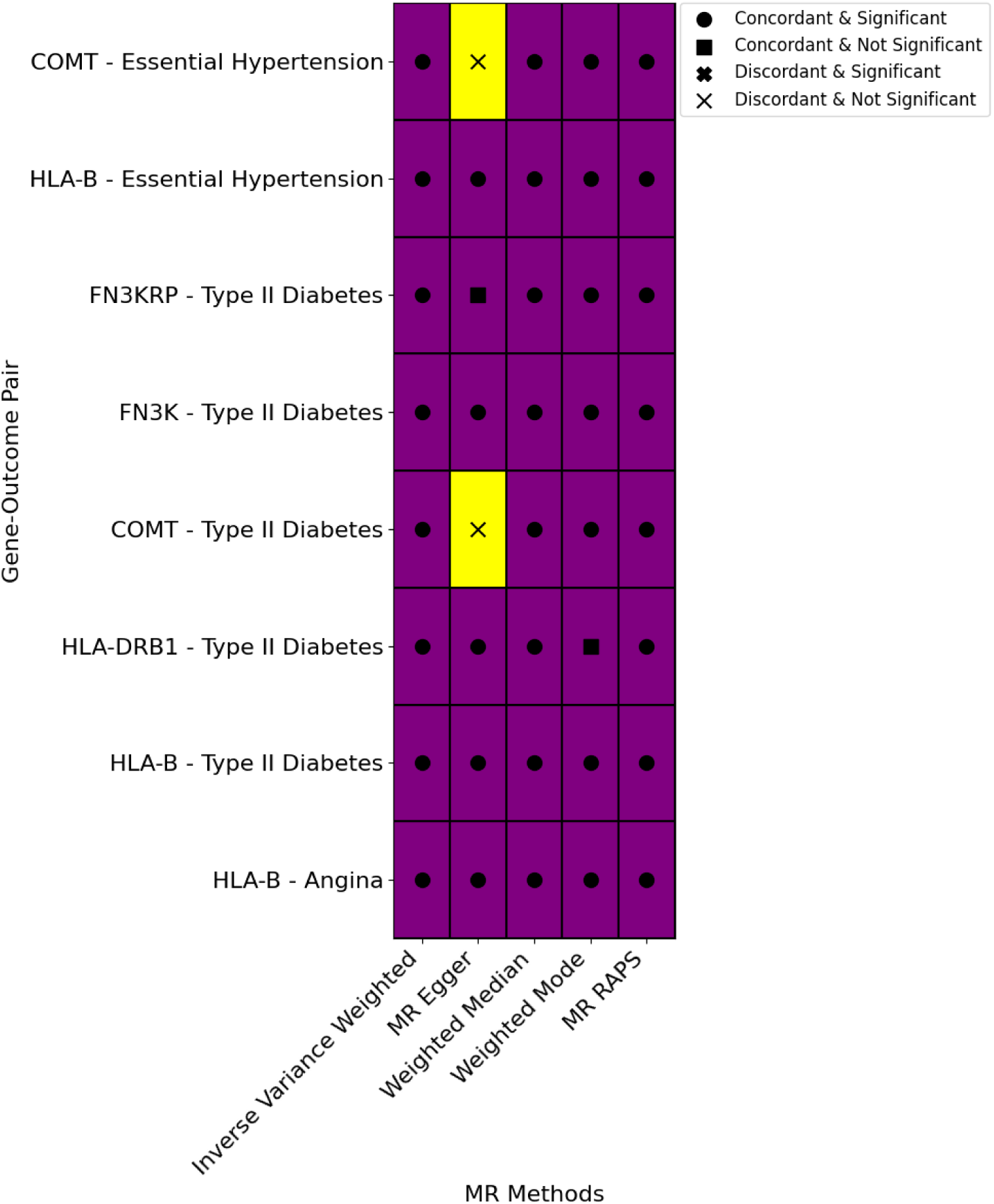
Heat-map of PBMC MR results, of those found to be significant in GSMR, compared with other methods. Each row in this figure represents a gene-outcome pairing, taken from those which were found to be significant in the GSMR results. Each column represents the result from a different MR method. Results are encoded either with: a filled in circle to represent a concordant and significant result, with that from GSMR, a filled square for a concordant and not significant result, a filled cross for a disconcordant and significant result and a light cross for a disconcordant and not significant result. Concordant results are shaded in purple and disconcordant results in yellow.

**Supplementary Figure 7:**
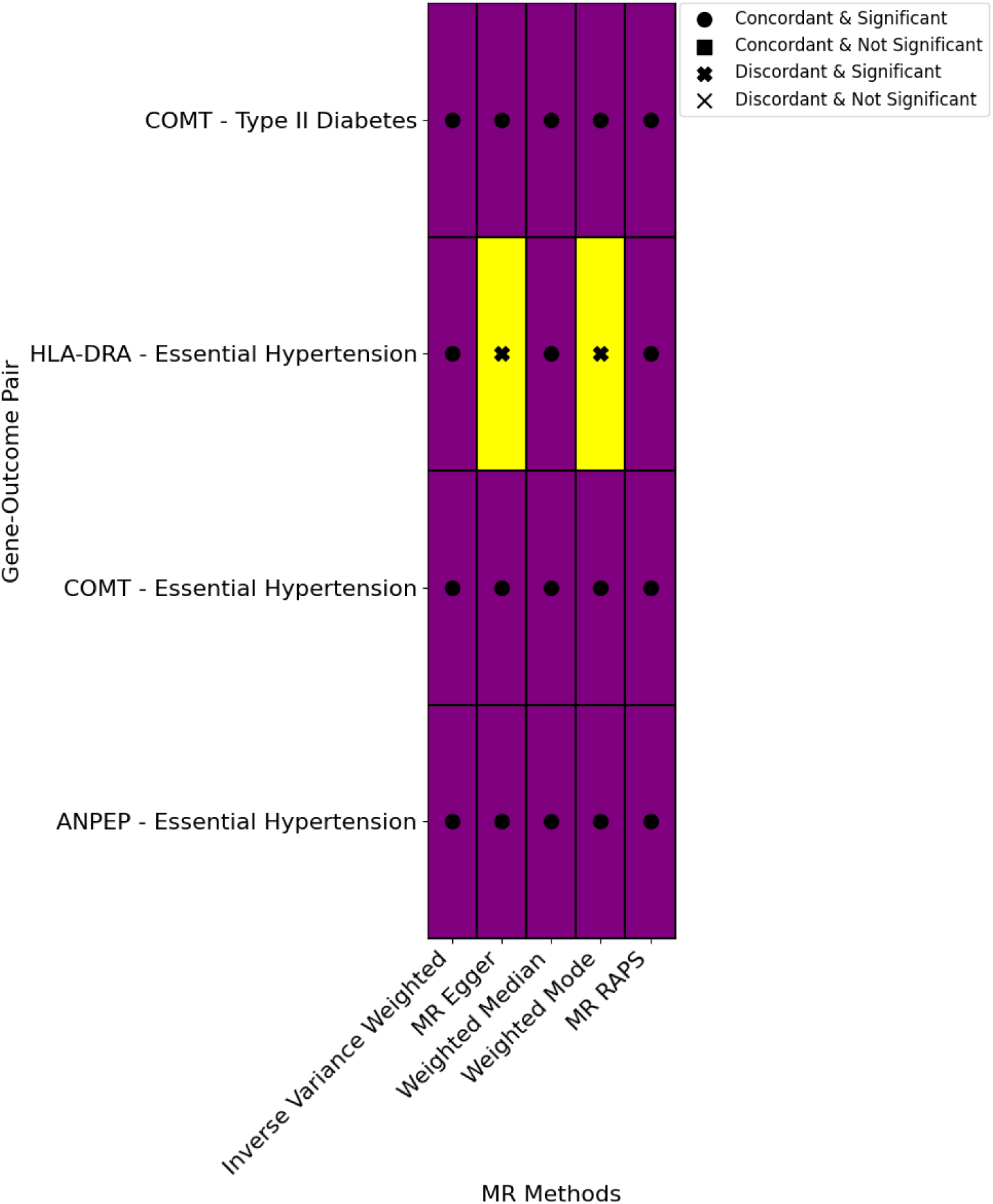
Heat-map comparison of MR results across methods for selected plasma protein–outcome pairs. This figure displays results from multiple Mendelian Randomisation (MR) methods for gene–outcome pairs that were identified as significant in the GSMR plasma analysis, and also testable in the GSMR PBMC analysis. These have been included for illustrative purposes, to demonstrate how results compare across different MR methods when robust GSMR associations are seen in both tissues. Each row in this figure represents a gene-outcome pairing, taken from those which were found to be significant in the plasma GSMR results and testable in the PBMC GSMR analysis. Each column represents the result from a different MR method. Results are encoded either with: a filled in circle to represent a concordant and significant result, with that from GSMR, a filled square for a concordant and not significant result, a filled cross for a discordant and significant result and a light cross for a discordant and not significant result. Concordant results are shaded in purple and discordant results in yellow.

#### Supplementary Figures 8-9

To assess whether differences in disease prevalence between the proteomics cohorts, Generation Scotland PBMC proteomics and UK Biobank plasma proteomics, or incidence in the outcome GWAS data bias the GSMR results, we performed sensitivity analyses for the two robust associations observed (COMT with type II diabetes and COMT with essential hypertension).

For each trait, we created modified GWAS datasets by randomly down-sampling cases from the UK Biobank exposure and outcome sets to empirically demonstrate that our MR results are robust to differences in prevalence between the Generation Scotland PBMC proteomics cohort and the UK Biobank plasma cohort, and to changes in incidence in the UK Biobank outcome cohort. For each resampled dataset, using the UK Biobank proteomics and outcome cohorts, we re-ran the SNP–exposure GWAS, the SNP–outcome GWAS (using REGENIE with Firth correction, which is robust to unbalanced case–control ratios [51–53]), and then repeated the GSMR analysis.

Figures 8 and 9 show the resulting GSMR estimates across these incidence scenarios. In both analyses, the estimated effect sizes remain stable (≈ −0.08 for type II diabetes, ≈ −0.06 for essential hypertension), with significance maintained once the outcome incidence exceeded modest thresholds. This indicates that the observed GSMR results are not sensitive to differences in prevalence/incidence across the cohorts.

**Supplementary Figure 8:**
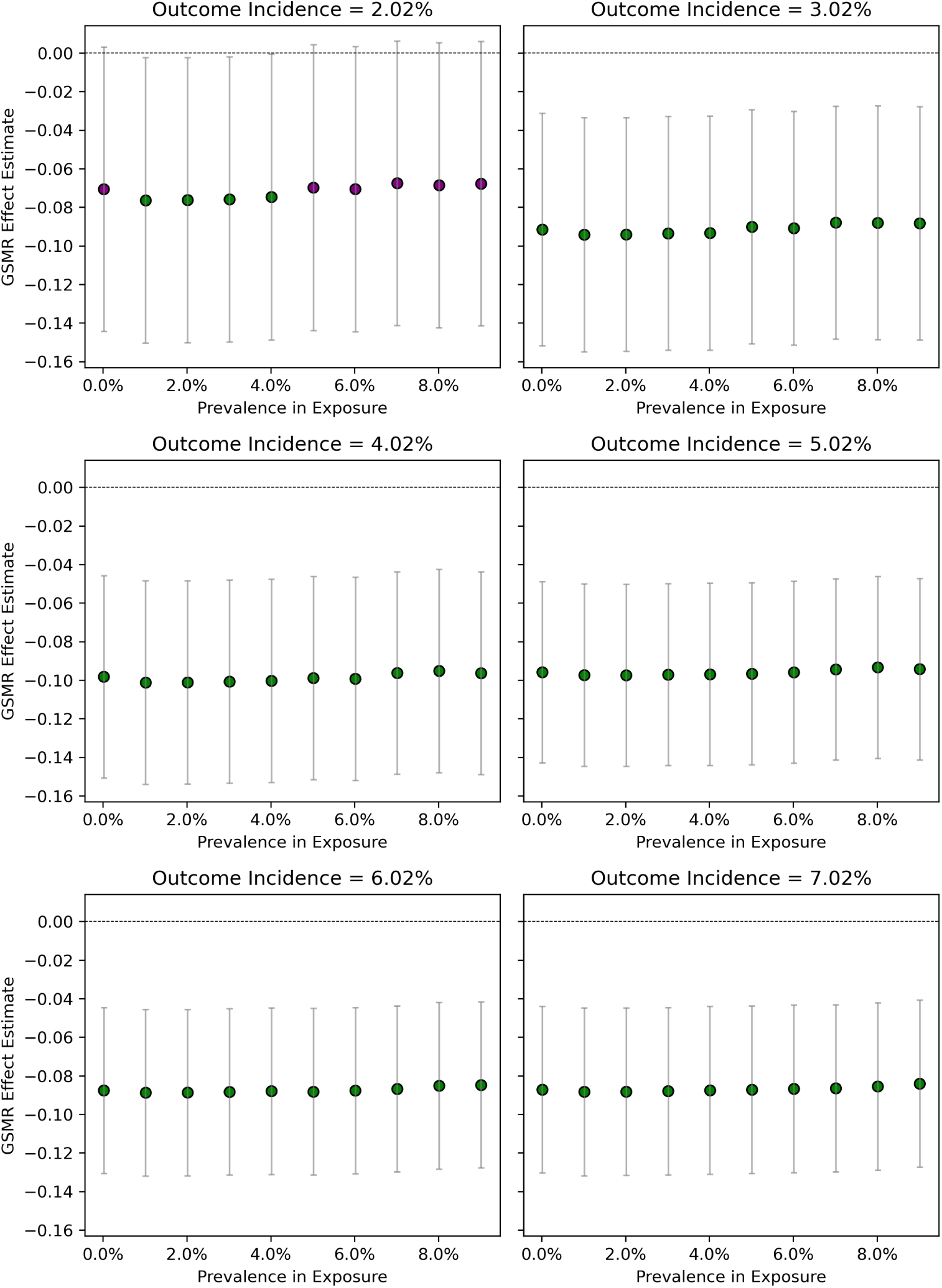
Sensitivity analyses for the COMT to E11 (type II diabetes) GSMR result. UKB exposure and outcome GWAS datasets were generated by randomly downsampling cases to create a range of disease prevalences/incidences (2.0% to 7.0%), approximating the values observed in the Generation Scotland PBMC proteomics cohort and the UK Biobank plasma proteomics cohorts. Each sub-plot shows the GSMR estimate re-calculated from resampled SNP–exposure and SNP–outcome GWAS. Points are coloured by significance (green = *P <* 0.05, purple = *P* ≥ 0.05). Error bars show 95% confidence intervals. Results stabilise once incidence exceeds ∼3%, with effect sizes consistently around –0.08, matching the main text.

**Supplementary Figure 9:**
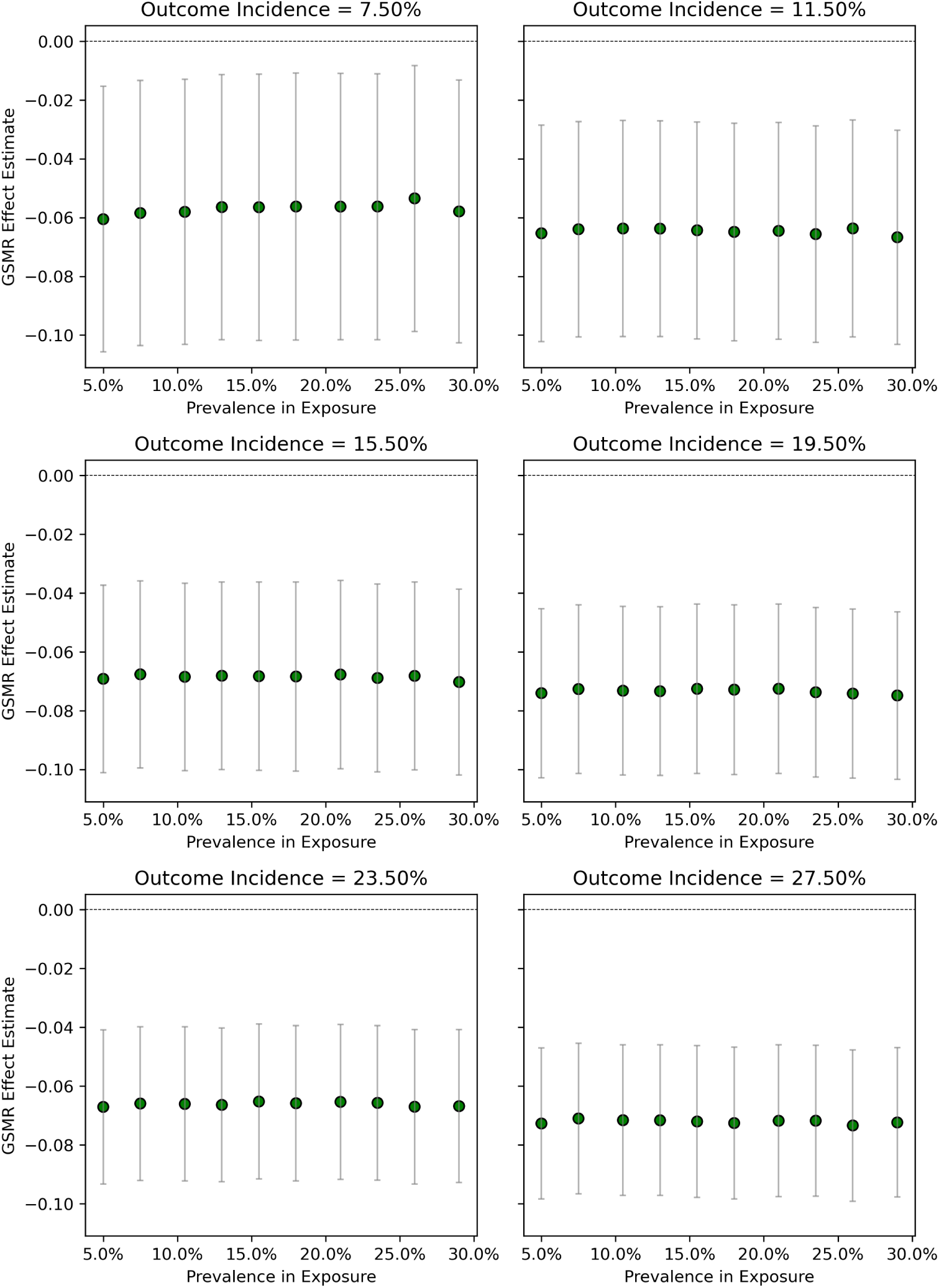
Sensitivity analyses for the COMT to I10 (essential hypertension) GSMR result. UKB exposure and outcome GWAS datasets were generated by randomly down-sampling cases to create a range of disease prevalences/incidences (7.5% to 27.5%), approximating the values observed in the Generation Scotland PBMC proteomics cohort and the UK Biobank plasma proteomics cohorts. Each sub-plot shows theGSMR estimate re-calculated from resampled SNP–exposure and SNP–outcome GWAS. Points are coloured by significance (green = *P <* 0.05). Error bars show 95% confidence intervals. Results remain stable across all incidence levels, with effect sizes consistently around –0.06, as reported in the main text.

#### Supplementary Information 1: IceR Parameters

IceR was run with the following parameters: min mz window = 0.001; min RT window = 1; feature mass deviation collapse = 0.002; only unmodified peptides = FALSE; align unknown = FALSE; use isotope peaks = TRUE; peak detection = TRUE; abundance estimation correction = TRUE; alignment score cut = 0.05; Quant pVal cut = 0.05; RT correction = TRUE; mz correction = TRUE; add PMPs = FALSE; plot peak detection = FALSE; calc protein LFQ = TRUE; kde resolution = 50; num peaks store = 5; MassSpec mode = Orbitrap; use IM data = FALSE. Feature level output from IceR was used for subsequent analyses.

## Notes

### Competing Interest Statement

The authors have declared no competing interest.

### Author Declarations

Generation Scotland participants provided written informed consent. Generation Scotland was granted Research Tissue Bank status by the East of Scotland Research Ethics Service committee (REC reference 15/ES/0040, 25/ES/0013). This project was approved by Generation Scotland as reference GS18318. The data/material transfer agreement was made on 05 Feb 2019. Researchers on this project had access to genotype data and pseudo-anonymised research data and materials. UK Biobank has approval from the North West Multi-centre Research Ethics Committee (REC reference 21/NW/0157; renewal of the original REC reference 11/NW/0382). Before being enrolled in the UK Biobank study, all participants provided written consent after being fully informed, adhering to the principles outlined in the Declaration of Helsinki. This research has been conducted using the UK Biobank Resource under Application Number 91924. The data and material transfer agreement was active from 21 Dec 2022. Researchers on this project had access to genotype data and pseudo-anonymised medical and biological data.

### Summary of Updates

We have repeated our analyses using outcome genome-wide association analyses that did not overlap those with measured proteomics data in UK Biobank. Similarly, we have refined our phenotype definition, and now use ICD10-measured outcomes. Our key results have remained largely unchanged, and we have now evaluated the robustness of our results by performing sensitivity analyses; we performed the same analysis with five other popular competitors to GSMR and obtain good concordance amongst the six mendelian randomisation estimation methods. Finally, we have corrected an error in the stated direction-of-effect of the GSMR results in the UK Biobank only analyses (that is, exposure and outcome genome-wide association study performed in UK Biobank).

